# Cohort profile: The I AM Frontier prospective cohort study in Flanders

**DOI:** 10.1101/2024.05.17.24307470

**Authors:** D Heylen, C. De Clerck, M. Pusparum, A. Correa Rojo, R. Van Den Heuvel, G. Baggerman, A. Standaert, J. Theunis, J. Hooyberghs, G. Ertaylan, N. Lambrechts

## Abstract

**Purpose:** The I AM Frontier cohort was set up to support proof-of-concepts aimed at precision health and more specifically personalized prevention and health promotion. The study was designed to identify patterns, markers and processes, that play a role in the spectrum between health and early onset of disease and may provide actionable information in a clinical setting, taking into account all ethical, legal and logistical aspects.

**Participants:** The first phase of the I AM Frontier study ran for 12 months as a longitudinal small-scale cohort study (n=30) in the Antwerp region of Flanders, Belgium. Participants were employees of the company hosting the study, they did not have a clinical diagnosis and were between 45-60 years old.

**Findings to date:** Even though no severe health problems are recorded at baseline, participants did report several physical complaints. There is a clear difference in longitudinal variation between clinical and research grade omics types, which might affect their respective ability to detect intermediate molecular changes that can be linked to phenotype changes.

**Future plans:** This cohort is being used to further support the design and implementation of a larger population health cohort with selected modalities for investigating feasibility of personalized prevention in real life setting. Future research will build on this longitudinal dataset to derive healthy yearly fluctuations (or normal ranges) at individual level for predicting early on-set deviations

**Registration:** The study was approved by the ethical committee of the Antwerp University Hospital (RegN°:B300201837314).

**Strengths and limitations summary:**

- The I AM Frontier proof-of-concept (POC) cohort study is unique in that it collected an extensive range of samples, with high longitudinal frequency, of healthy individuals for 12 months. The implemented sampling technologies (for clinical parameters, whole genome sequencing (WGS), methylation, quantitative proteomics, metabolomics, microbiome, retina scans, wearables, and standardized questionnaires on e.g. food intake and medical status in combination with genome sequencing at the start of the study) were selected to maximize overlap with large cross-sectional studies and biobanks such as e.g. UK biobank to allow comparison of phenotypical profiles present across different studies.
- The highly granular (i.e. collected with high longitudinal frequency) data within this study allows us to construct dense participant profiles. Frequent longitudinal data collection of multi-omics data is emerging with new technical advancements for the in-depth analyses of molecules in small blood volumes. To allow the routine usage of such measurements in clinical practice, the temporal changes observed in this cohort can serve to evaluate the frequency and added value of such highly granular measurements.
- Perceptions of precision health, such as communication of clinical follow-up data, personal risks from genomics, behavioral aspects, and the ethical dilemmas that go together with all of this, are included in the scope of the cohort.

## Introduction: A healthcare transition towards personalized prevention

The healthcare landscape is evolving, and several trends push the current socio-technical regime beyond its (financial) capacity. With an aging population, the prevalence of chronic diseases such as diabetes, cardiovascular diseases, and cancer is increasing, along with disease co-morbidity. The European Union Policy Forum estimates that about 70-80% of the total healthcare cost in the EU (about 700 billion Euros) is earmarked for chronic care. Furthermore, the WHO predicts that partially preventable diseases will constitute the main contribution to the burden of disease in 2030. The indirect costs, including loss of income, inability to work, and informal care, are a substantial additional cost to society (1,2).

On top of the increasing financial and social costs, there is growing scientific consensus that conventional approaches towards disease and therapy are not generating sufficient new solutions for complex diseases, and that health promotion and data-driven disease prevention are the way forward (3,4). Therefore, a reshaping of the socio-technical regime is imperative. Society must evolve towards a more sustainable healthcare system. In that light, the concept of ‘precision health’ is introduced, focusing on a holistic approach to healthcare, maintaining and improving health by encompassing a wide range of factors influencing health, such as behavior and environmental factors. As science understands more about a person’s biological makeup, it becomes clear that many causes of disease and causes of unresponsiveness or serious side effects to treatment can be explained by the variability in an individual’s characteristics. Precision health tackles these problems by moving away from a ‘one size fits all’ approach by integrating data on the dynamic biological makeup of individuals, and the lifestyle and environmental factors that interact with this makeup, to develop a complex and individual phenotype (5,6).

So-called ‘personalized prevention,’ on the other hand, is a specific aspect of precision health that envisions a personal approach for disease prevention and health promotion (7).

Recent technological advancements, such as personal sensor technologies, and developments in various ‘omics’ fields, such as genomics, proteomics, and transcriptomics, increase the potential of precision health by improving the comprehension of an individual’s physiological state. Understanding the integrations of underlying biological processes involved in disease onset and development, rather than looking at isolated parameters of these biological processes, provides opportunities for increased prevention, better diagnostics and ultimately more efficient treatment. The integrated Personal Omic Profiling (iPOP) system by Michael Snyder in 2012 (8) is an analysis methodology that combines multi-omics information with monitoring of physiological state. Studies following this nascent effort have demonstrated that longitudinal iPOP can be used to interpret healthy and diseased states (9,10) .

### Objectives and aims of the I AM Frontier Study

Today, various biobanks and cohorts collect samples and perform in depth phenotyping of individuals with the purpose of facilitating precision health across various biomedical domains (9,11–15). However, data collection in a cohort with healthy individuals by integrating extensive omics, clinical and continuous personal monitoring data, is still scarce. With the insights from a cohort study of this nature, we aim to provide insights into the earliest manifestation of chronic diseases and enable tracking of disease progression. The I AM frontier proof-of-concept (POC) study was set up as a first step towards larger scale-ups by gaining specific knowledge on the added clinical value of high frequent tracking of a generally healthy cohort of individuals with clinical tests, omics tests, surveys, and wearables while considering ethical and legal aspects. The study investigates candidate processes (or molecules) tightly regulated in health (considering seasonality) and their deviation from their normal pattern, as this could signal the onset of a spectrum of diseases.

Besides the clinical aspects, we’ve also considered the ethical and societal impact by studying best practice on how to communicate gained insights to individuals informing them about potential health threats whilst avoiding ungrounded life-disrupting messages.

### Cohort description

The first phase of the I AM Frontier study ran for 12 months, starting in March 2019, as a proof-of-concept cohort study aimed at advancing personalized disease prevention (n=30) in the Antwerp region of Flanders, Belgium (Figure 1). The cohort includes comprehensive longitudinal multi-omics, clinical, and sensor-based health monitoring data. This, together with wide-ranging questionnaires, creates detailed phenotyping of healthy individuals.

**Figure 1:**
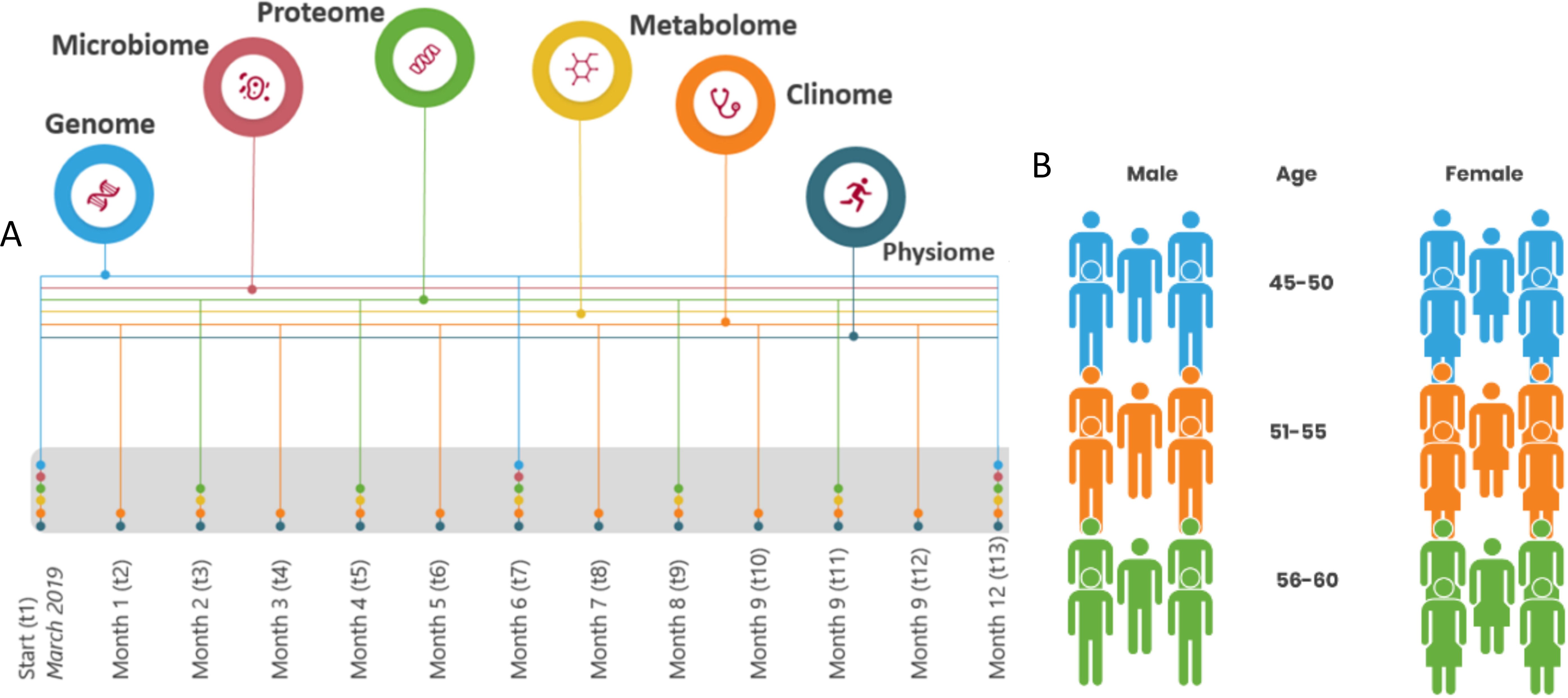
A) Data collection flow diagram. Datatypes were collected at different time points, circle stacks on each time point indicate which datatypes are collected at that moment in time (n). B) Gender and age distribution of the cohort. Cohort consists of 30 participants (n=30).

Besides the scientific results, also logistical and ethical know-how was obtained with the cohort. The logistic results focus on how the study collected, stored, and analyzed various samples and data types (blood, urine, hair, saliva, stool, retina scans), either monthly or (for some) quarterly. Various steps in the processes had different sensitivities, processing times, and operating costs. Logistical processes and workflows were created and reported to enable future optimization efforts when scaling up similar cohort studies. Ethical insights were gained by interviewing participants regarding their perspective on personal health data and communication on its insights.

### Patient and public involvement: study design and eligibility criteria

The IAM frontier cohort study exclusively targeted its own Flemish research institute’s employees, with the expectation that, due to the nature of their employer, employees would be more open to research-grade technologies. With an intensive protocol, the nature of this POC cohort requires sufficient commitment and understanding of what is asked and of the risks and uncertainty of what may be found. As this study is a POC, representativeness is not a requisite; feasibility, on the other hand, is. Other reasons for this target population are also of a practical nature, i.e., the occupational health service did sample collection during working hours. Privacy and avoidance of any pressure to participate or remain in the study were given highest priority. The age range of 45-65 was chosen because it encompasses the period during which chronic diseases are most commonly initiated .

All 842 VITO employees were initially approached through a company-wide information session and the internal website. Subsequently, from the 270 interested candidates, 76 met the age requirement: an age range of 45-65 was chosen because it encompasses the period during which chronic diseases are most commonly initiated (16). These 76 candidates were contacted for an interview and received additional general information about the project. A set of exclusion criteria was used, rejecting people suffering from a chronic disease (i.e. diagnosed and currently followed-up by a medical specialist for asthma, chronic bronchitis, chronic obstructive pulmonary disease, emphysema, myocardial infarction, coronary heart disease (angina pectoris), other serious heart disease, stroke (cerebral haemorrhage, cerebral thrombosis), diabetes and cancer (malignant tumour, also including leukaemia and lymphoma)). The researchers ensured that none of them were frequent blood-donors (due to interference with sampling protocol), were active smartphone users (as some degree of digital literacy was required throughout the study), and that their motivation was not based on personal health gain but research-oriented. Based on these criteria, 66 of the 76 candidates could still be considered eligible. Next, we ranked them according to age and selected 30 participants with an equally spread age distribution in the 45-60 age range, and an equal amount of female and male participants (Figure 1B). The candidate selection process is shown in a flow diagram in supplementary figure 1. All 30 participants remained in the study throughout the first year of the sampling process. Only in the very last month of the sampling procedure we experienced ten drop outs due to the COVID-19 pandemic and lock-down that was in place during sampling. All participants signed an informed consent and the study was approved by the ethical committee of the Antwerp University Hospital (RegN°:B300201837314).

Supplementary figure 1: Candidate selection process

### Patient and public involvement: Feedback to the participants

Three different types of results were communicated to the participants. The data collected within the study can be divided into three groups based on the actionable nature of a datatype.

The first group consists of clinical parameters associated with a known adverse outcome. The participant received its values for the clinical parameters compared to the normal clinical ranges, provided by AML Flanders (17), through individual reports on a bi-monthly basis. The study doctor discussed these reports with the participants every four months and advised them on actions that could positively affect these parameters. As the project intended to measure numerous biological data points, including genomic data, so-called ‘incidental findings’ (IF), could be detected, the second feedback result. The IF refer to genetic alterations associated with conditions or diseases unrelated to the patient’s or participant’s current condition for which current tests are being performed. The ‘ACMG Recommendations for Reporting of Incidental Findings in Clinical Exome and Genome Sequencing’ (18,19) was followed for communication of IF or discovery findings based on IF. All other ‘omics’ parameters are considered in the final group and were kept for scientific purposed only, as they were associated with insufficient evidence-base to an adverse outcome in scientific literature. These results were therefore only used for research purposes and were not communicated to neither the study doctor, nor the participants.

### Study sampling and data types

Sampling of the first phase took place between March 2019 and March 2020 and included the collection of different human biospecimen (blood, urine, saliva, stool, and hair) and monitoring data (wearables, questionnaires, and retina scans). See Table 1 for an overview of all collected data, the collection time frames, the measurement techniques and specific laboratories in which the different analyses were performed within the I AM Frontier POC study.

**Table 1.**
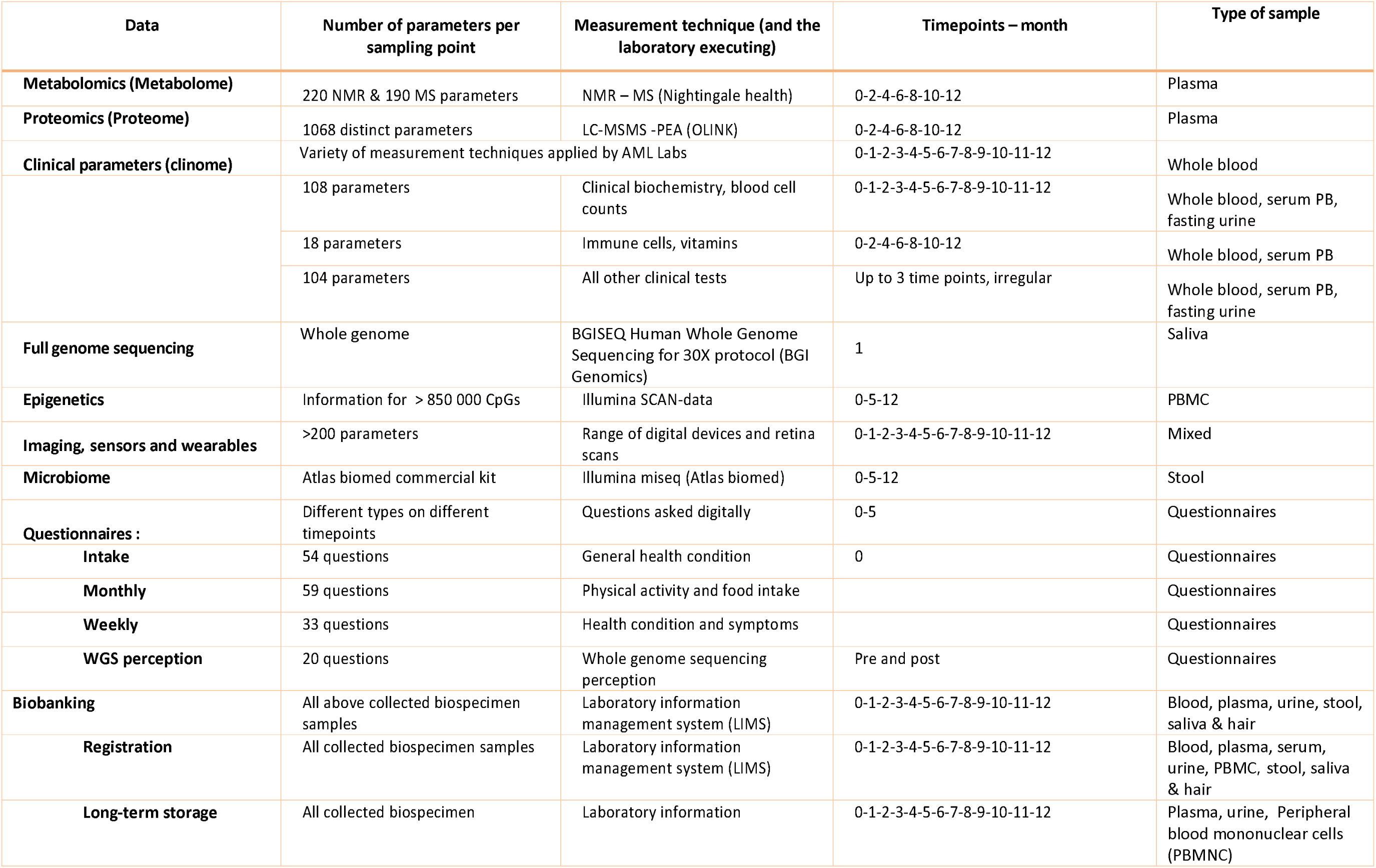
Visual of the data collection for I AM Frontier Cohort. A parameter specific overview of the measured variables for metabolomics, proteomics and clinical parameters can be found in supplementary tables (2–4).

Monthly appointments were made with the participants for a health examination survey and bio-sample collection. Sampling was done by a nurse and under the supervision of an occupational physician at the in-house medical department. Blood pressure, body height, weight, and abdominal circumference were measured in a standardized way by the physician.

Blood was drawn by a nurse in the morning after a minimum of 8 hours of fasting. Multiple blood samples were collected in EDTA-, citrate- and serum- vacutainers (Greiner Bio-One International, Vilvoorde, Belgium). Blood was stored at room temperature till further processing. Serum was collected by centrifugation at 3500 rpm for 10 min. EDTA-whole blood, citrate whole blood, and serum were transported (at room temperature) to the clinical laboratory within 6 hours after collection for further processing and analysis. Plasma was removed by centrifugation at 3500 rpm for 5 min. Aliquots were made in pre-cooled tubes and immediately stored at -80°C. LeucosepTM tubes (Greiner Bio-One International, Vilvoorde, Belgium) were used to isolate peripheral blood mononuclear cells (PBMC) from EDTA-blood (30 min, 400xg). The PBMC suspension was divided into two aliquots (each originating from 5 mL of the original whole blood sample) and spun down (10min, 250xg) to create PBMC pellets, which were stored at -80°C till the DNA extraction. Genomic DNA (gDNA) was extracted from PBMC pellets using QIAamp DNA Blood Mini Kit (Qiagen, Antwerp, Belgium) according to the manufacturer’s protocol.

Urine samples were taken after an overnight fast and stored immediately at 4°C. Aliquots were stored at -80°C. Saliva samples were collected using Oragene OG-500 self-sampling kits (DNA Genotek, Ottawa, Canada). Stool samples were collected at home using Atlas Microbiome Tests (Atlas Biomed, London, UK). Saliva and stool samples were stored at room temperature till shipment for further analysis. A lock of hair with a thickness of a match (2 mm) and a length of 4 cm was taken and stored in an envelope at room temperature.

All biospecimen samples were registered at the Biobank@VITO (Mol, Belgium, ID:BB190064) and either stored temporally in the VITO’s biobank facilities until transfer to laboratories for analysis or stored long-term in the VITO’s biobank facilities. Standard operation procedures (SOPs) to collect and process blood, urine, saliva, stool and hair samples were implemented. All samples have been collected in accordance with the applicable Belgian regulations regarding the use of human body material for scientific research (Belgian Law on use of human body material, 2008) and the Belgian Royal Decree on biobanks (Het Koninklijk Besluit betreffende de biobanken. Belgisch Staatsblad 05.02.2018. Brussels (20)).

For the digital monitoring data all participants received a Garmin Vivosmart 4 activity watch to record their heart rate, activity, and sleep on a continuous basis to have accurate readings. Retinal imaging was performed at the sample collection visits using a Canon CR-2 non-mydriatic digital retinal camera with CE mark in accordance with European regulations (Hospithera, Brussels, Belgium). The method of De Boever and colleagues was used (21,22). Survey monitoring data was gathered by participants completing self-administered questionnaires to assess their personal lifestyle, environment, and health status. A questionnaire at the start of the study was used to set the intake status of the participant, such as information on previous or ongoing diseases. Each month, the day before sampling, participants completed questionnaires on daily activity and lifestyle patterns (nutrition, smoking, etc.). Additionally, a weekly questionnaire, based on the Cohen-Hoberman Inventory of Physical Symptoms (CHIPS) method, was used to inquire health and well-being in the last seven days (23). This methodology consists of 33 questions for which the participant had to estimate how much a specific disorder had bothered them over the previous seven days on a scale from 0 (painless) to 4 (severe pain). Finally, participant’s perspective regarding ethical communication of insights derived from sampled health data and feedback on their participation in the study was questioned. Questionnaires were sent and completed digitally. The questions and respective answers were coded based on a codebook. The coded data were saved directly in a data platform and could only be accessed by the involved researchers.

### Data Analysis Techniques

To correct for non-biological variance within the data, systematical data pre-processing was performed as prescribed by the laboratories performing the data measurement.

An initial analysis of the data obtained from the diverse sampling technologies mentioned above, was conducted to formulate the information presented in the ‘Findings to Data’ section. The methodology of Allen et. al (23) was followed to analyze the CHIPS questionnaire survey results. This study reported that the 33 survey questions concerning health complaints can be grouped into eight clusters. The identified physical health clusters are Sympathetic/cardiac symptoms, Muscular pain, Metabolic symptoms, Gastrointestinal symptoms, Vasovagal symptoms, Cold/flu, and Minor hemorrhagic symptoms.

The clinical results are summarized in some key clinical parameters, such as body mass index (BMI), Framingham risk score for cardiovascular disease (24), and the ADA diabetes risk score (25). Clinical parameters were measured either each month, every other month, or only thrice during the 12-month period. Each of the monthly clinical parameters are grouped in clinical health clusters: Adrenal, Allergy, Anemia, Carbohydrate metabolism, Cardiovascular, Hormonal, Immunological, Inflammation, Ionagram, Kidney, Liver, Proteins, and Thrombosis. The clinical health clusters were determined in consultation with the study physician, grouping together clinical parameters to simplify communication with participants. Measurements of the clinical parameters are compared to the normal interval for this parameter based on (AML) medical laboratory reference values (17) (26).

### Findings to date

As this was the first phase of the I AM Cohort POC, we report the main findings from the first clinical and omics analytics.

### Survey results

In the cohort, physical symptoms were self-reported with the CHIPS methodology. Supplementary figure 2) shows that at the start of the cohort study, more than 50% of the participants had sleeping problems and back pain. 20% of the cohort indicate their sleeping problems to be serious. In addition, more than 30% of the participants suffered from constant fatigue, tense muscles, headache, muscle pain, and low energy at the start of the study (Suppl. figure 2).

Supplementary figure 2: Baseline physical symptoms of IAM Frontier participants obtained at the time of recruitment with the CHIPS methodology from 0 (in green: the least severe/painless) to 4 (in red: severe pain). The horizontal width of each colored bar indicates how many participants indicate to suffer from that specific physical symptom with the corresponding severity.

To highlight the longitudinal aspect of physical symptoms in the cohort, Figure 2 shows an overview of the self-reported physical symptoms for both men (Figure 2A) and women (Figure 2B) throughout the first phase sampling. The color indicates the severity of the complaint, the more intense the color the more a participant felt its impact. Each question falls within one of eight physical health clusters, indicated on the right of the figure (23). An interesting lead provided by the Figure 2 is that the physical health cluster with the most prevalent reported symptoms is the cluster for metabolic symptoms, with sleep problems as one of the most reported complaints. Additionally, it might be interesting to explore the higher prevalence of self-reported headaches by the female population compared to the male population. Over the longitudinal course of the study, most physical health clusters show high internal consistency, as participants report similar levels of severity for symptoms within one cluster. For certain specific symptoms participants did experience fluctuations in severity. This was the case for indigestion, muscle pain, headache, sleeping problems, and back pain. Finally, symptoms of the Cold/Flu cluster were reported less during the summer months, as expected, with Cold/Flu season situated during the winter months (Figure 2).

**Figure 2:**
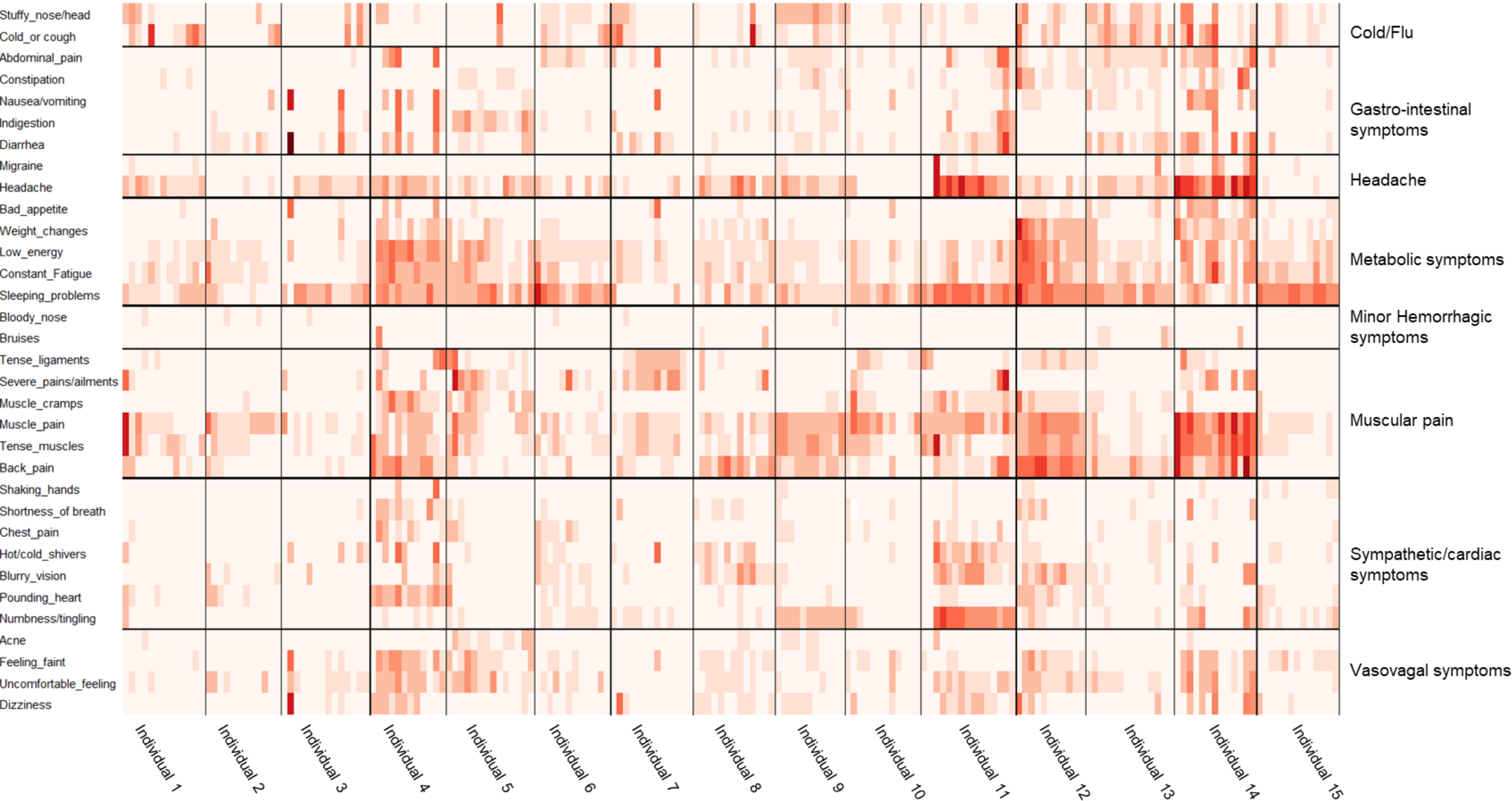
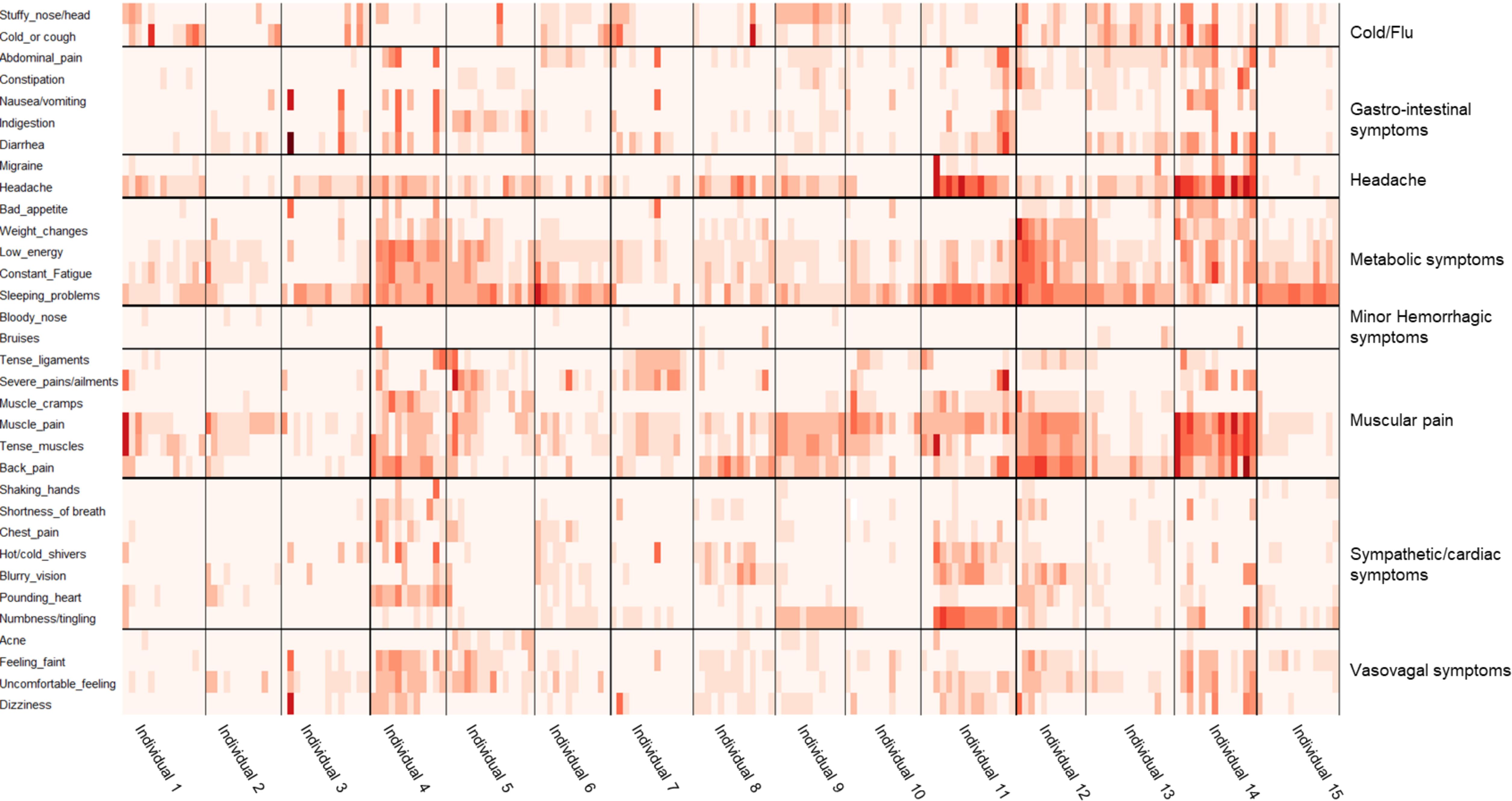
Physical symptoms throughout the study period (timepoints 0 to 12) were obtained via questionnaires and grouped according to the findings of Allen et. al (23) . The color intensity indicates the severity of the complaint on a scale from zero to four. Month zero is March 2019, and month 12 is March 2020. A) Male participants all time points. B) Female participants all time points.

### Clinical results

Similar to the survey-reported physical symptoms findings, a similar overview can be provided for the clinical measurement findings. At the start of the study, the body mass index (BMI) ranged from 19 to 34 kg/m2, with a mean of 25 kg/m2. On average, the BMI for men was significantly higher than for women. Also, the average blood pressure was lower in women (120/80 mmHg) than in men (129/84 mmHg). Only one person was identified as a current smoker, while the percentage of former smokers was lower in women (13%) than in men (27%). On average, the Framingham risk score for cardiovascular disease was 0.6 for women and 5.9 for men, corresponding to a 0.6% and 5.9% 10-year risk of a first ‘hard’ cardiovascular event (e.g. heart attack), respectively. The mean of the ADA diabetes risk score was below 5% in both women and men, corresponding to a low risk of type II diabetes mellitus. The detailed descriptive statistics of these parameters and the other clinical biochemistry parameters, such as total cholesterol, HDL cholesterol, and glucose, are reported in table 2.

**Table 2:**
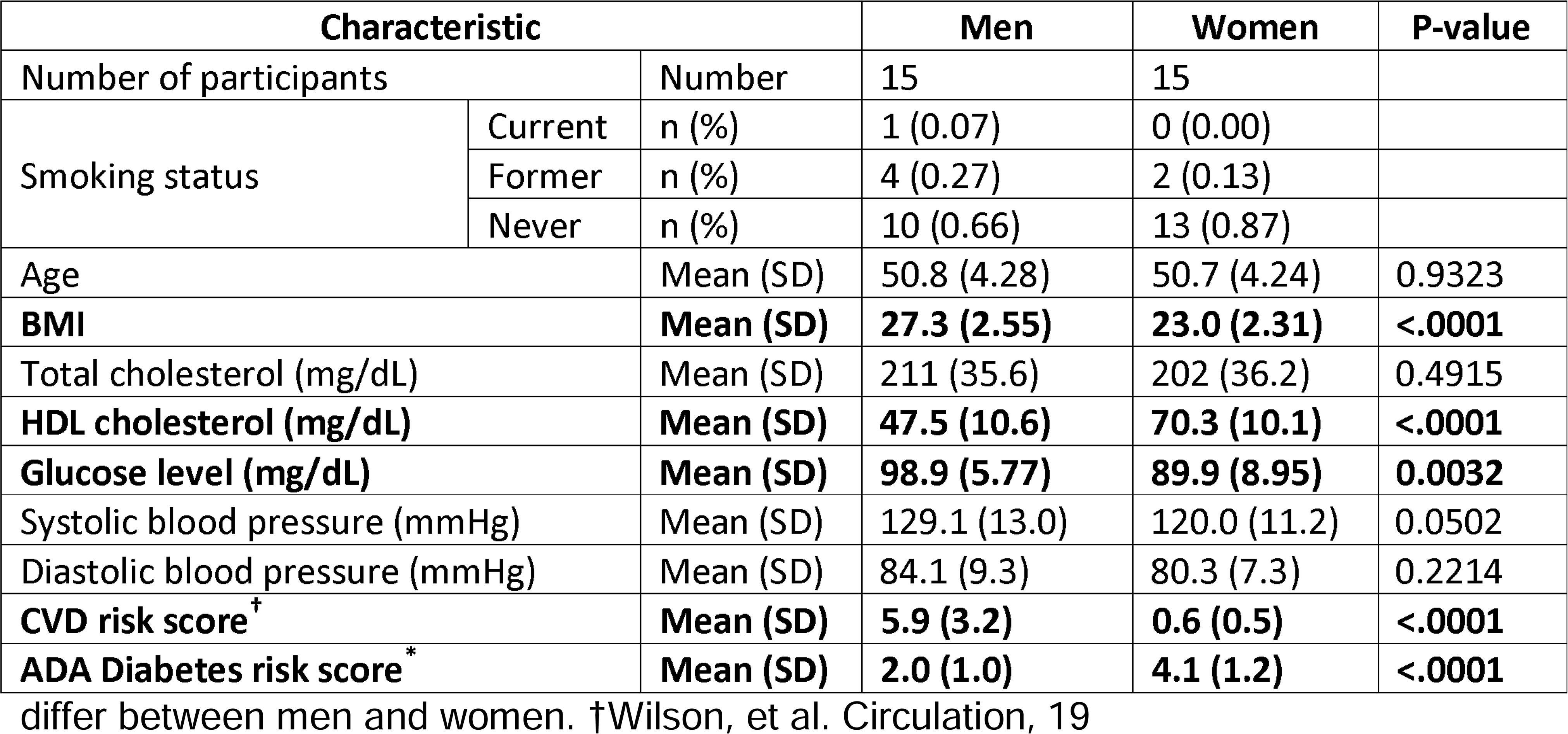
Baseline characteristics at the time of recruitment of IAM Frontier by gender. The Student’s t-test was performed to determine if the means of each characteristic significantly.

Figure 3 is a visual representation of the monthly measured clinical parameters for each participant during the one-year course of the study, grouped into clinical health clusters, indicated on the right axis of the figure. A red measurement means the clinical parameter was out of bound compared to the normal interval for this parameter. If the value was within normal bounds, it is represented in white. Values represented in grey indicate a missing value. Figure 3A, shows clinical parameters for the male population, Figure 3B for the female population.

**Figure 3A:**
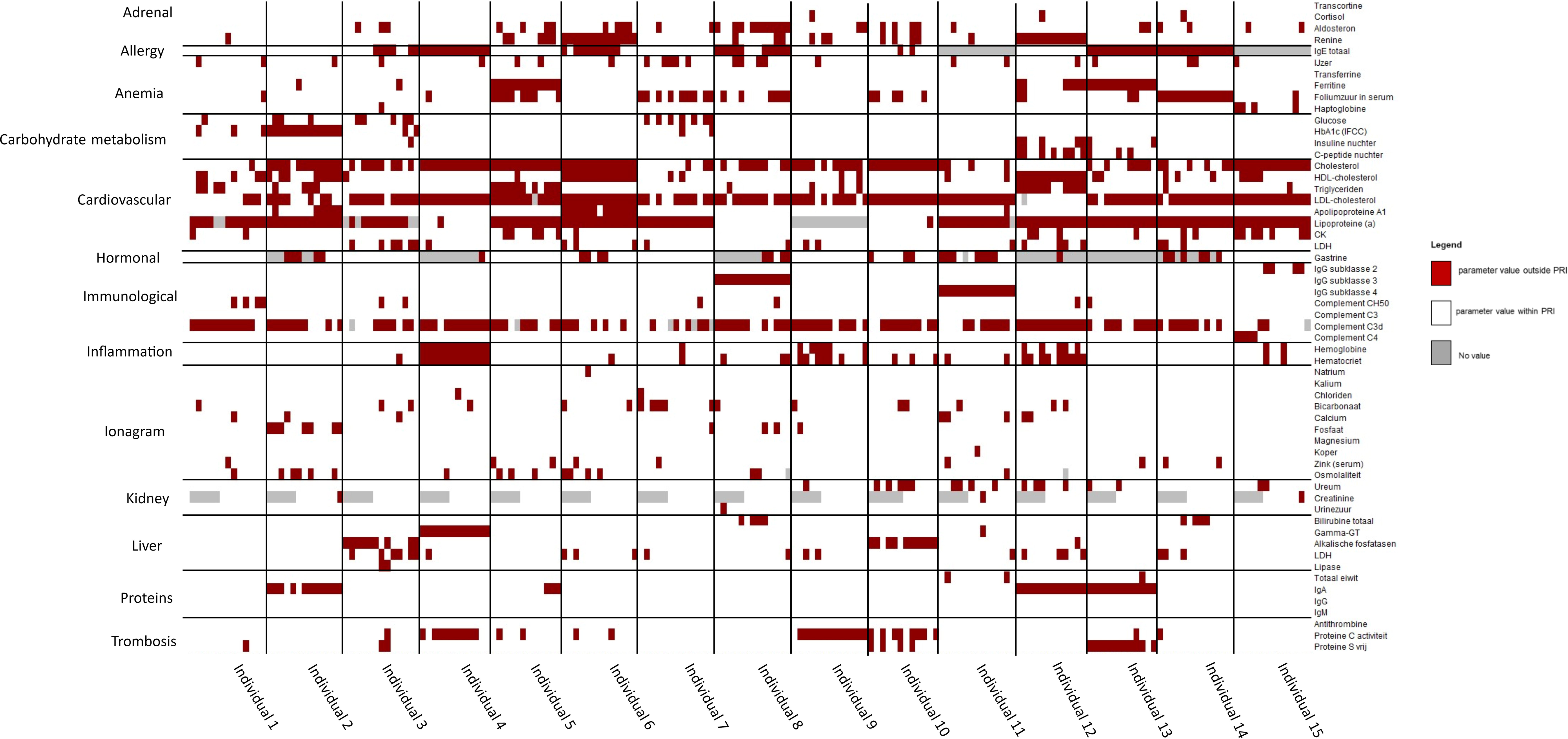
Longitudinal heatmap representing which clinical parameters (right), grouped in clinical health clusters (left), are out of normal range bounds, i.e. population reference intervals (PRI), for male participants.

**Figure 3B:**
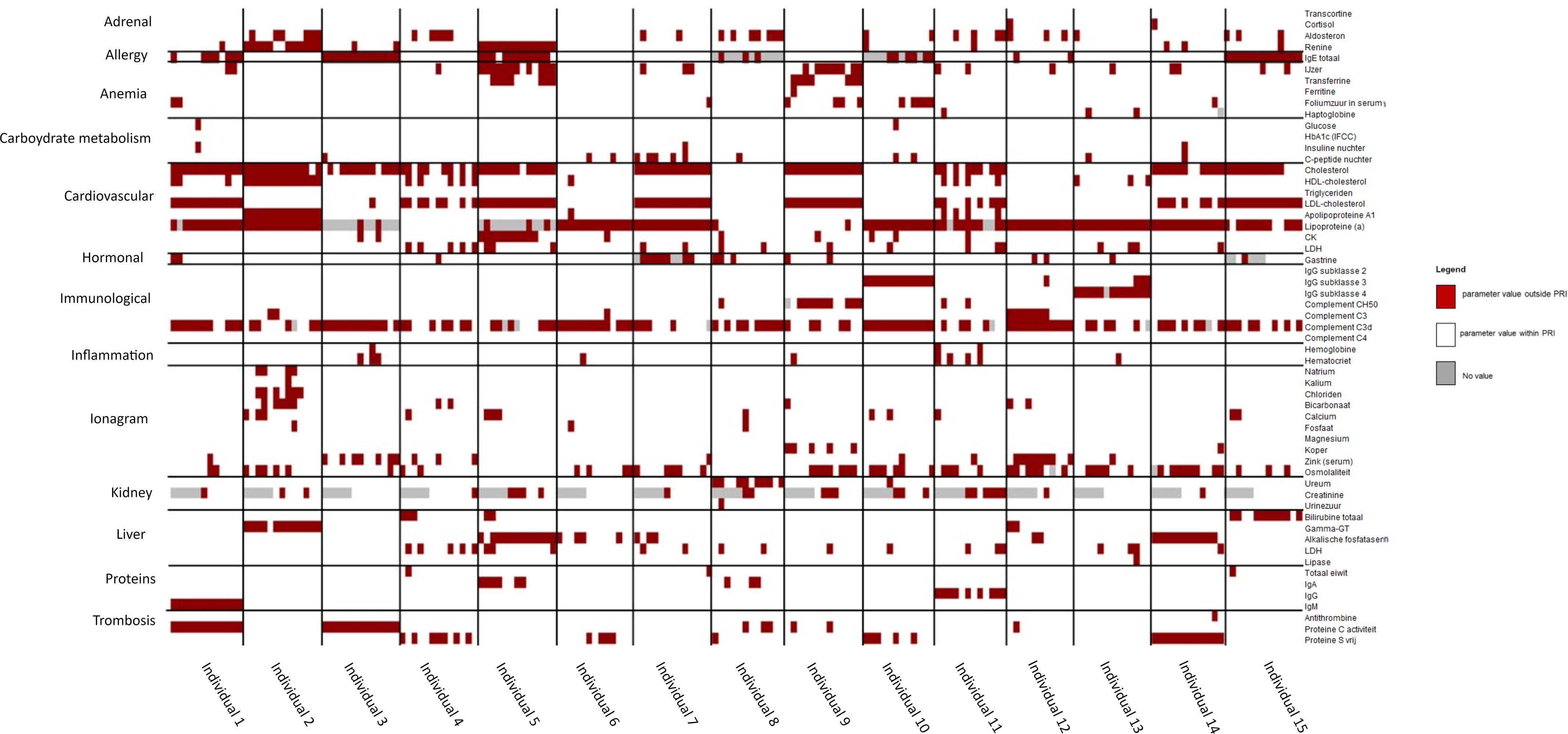
Longitudinal heatmap representing which clinical parameters (right), grouped in clinical health clusters (left), are out of normal range bounds, i.e. population reference intervals (PRI), for female participants.

All male and female participants had at least one clinical parameter that is out of bounds at any time. A limited number of participants showed an obvious change in the clinical parameters over time. One of the most striking patterns was the C3d complement parameter, which was out of bounds (increased) for every participant, at least at some point during the sampling period. High C3d complement levels point towards an increased activation of the immune response through activation of the complement system (27).

Besides that, we noticed an obvious difference in the cardiovascular parameters between men and women in the cohort. Especially for cholesterol & LDL-cholesterol, more aberrant measurements are measured for men than for women.

The detailed longitudinal visual exploration of the clinical data in Figure 3 was done to search for any (time-related) clinical events that coincide/can be related to the appearance of the physical disorders, as shown in Figure 2. Visually comparing the results of the clinical parameters (Figure 3) with the survey parameters (Figure 2) did not immediately yield obvious one-on-one interactions. Further research will have to examine if predicting a significant correlation between the self-reported symptoms, clinical and omics parameters is possible.

### Genetic results

Whole-genome sequencing was performed for all 30 participants to detect variants such as single-nucleotide polymorphisms (SNPs), and insertions and deletions (INDELs) (28) that could be associated with chronic diseases or illness indicators. On average, 3807610 SNPs and 855955 INDELs were identified for each participant. Of these variants, 94.43% were registered in the database for single nucleotide polymorphism dbSNP (build 138) (29). Out of all SNPs analyzed there were 11342 synonymous, 11597 missense, 168 stop gain, 61 stop loss, and 38 start loss mutations.

Next, the participants’ genome results were compared with the ACMG SF v2.0 list, a list of 59 genes for which it is considered as clinically evident that pathogenic variants of the genes result in diseases that might be prevented or treated. This resulted in two hits (19). One person had a pathogenic variant of the BRCA2 gene (rs28897756) related to breast cancer. The other person had mutations in the MUTYH gene (rs34612342) related to colorectal cancer. Communication of those genetic IFs towards the participants was handled according to the strategy described earlier in the feedback to participants’ section. Qualitative evaluation of this communication strategy will be researched in a next phase.

### Genetic background

For quality control, the genetic structure of the IAM Frontier cohort was compared with other populations worldwide (Suppl. Figure 3). A principal component analysis (PCA) was used for this endeavor, with the identified variants in the whole-genome sequences and the 1000 Genomes Project as a reference panel. Only SNPs located on autosomes with a minor allele frequency above 5% and a linkage disequilibrium of less than 0.3 were considered in the analysis. The first two components captured the main variance with 48.55% and 23.88%, respectively. Using these two components, the 30 participants of the IAM Frontier study were mapped as a European population with overlaps on Admixed populations, as seen in Suppl. Figure 3A.

PCA was repeated but only with European sub-populations in the 1000 Genomes Project using the same set of filtered SNPs as described above to see the differences between other European populations. Suppl. Figure 3B presents the results where most participants were mapped between the British and Scottish Population and Utah residents with European ancestry. These findings support the validity of our genomics data as all IAM Frontier participants were reported to have European backgrounds, based on the information obtained from the self-reported ethnic background questionnaire.

Supplementary figure 3: **A)** Genetic structure of I AM Frontier cohort as red points compared with worldwide populations, **B)** and with only European sub-populations.

### Multi-Omics results

Figure 4 gives some preliminary information on the diverse data collection and the longitudinal robustness of the cohort study participants’ clinical biochemistry, proteomics, and metabolomics measurements, respectively. The various targeted parameters represent the biomolecular entities of the collected bio-samples. It is generally accepted that these entities reside in the same cell environments and affect the same clinical endpoint/phenotype (30). However, Figure 4 shows that these somehow intertwined data layers have different longitudinal stability within and across individuals. For the clinical variables, measurements remain relatively constant over time compared to the protein and metabolite variables. Various data pre-processing steps and batch corrections were performed for the different data type (see data analysis techniques). Thus, we could rule out that this difference in longitudinal stability between different types of omics data was due to variations in the technical sampling process.

**Figure 4:**
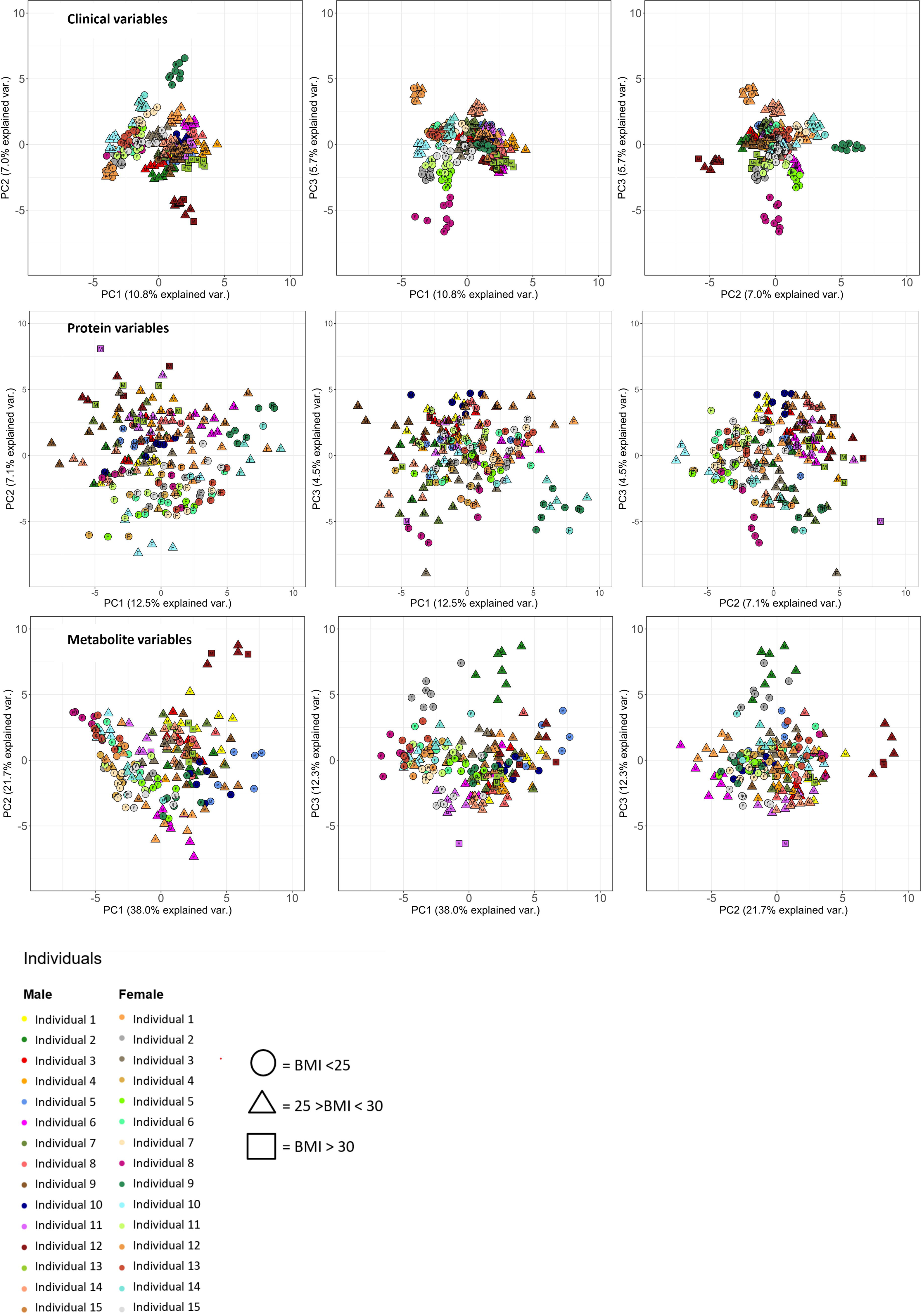
PCA plots of the various longitudinal omics datatypes. The three principle components that capture most of the variance in each data type are plotted, resulting in three possible combinations to plot these PC’s on a one-to-one basis. **A)** clinome data, **B)** proteomics, **C)** metabolomics, annotated with gender (M = Male, F = Female) and BMI information. Each point in the plot represents an individual at one specific time point, different measurements from a same individual are represented in one colour. The longitudinal character of the I AM Frontier data reveals that clinical parameters behave more stable over time and produce more dense inter-personal clusters compared to proteomics and metabolomics data.

## Discussion

The I AM Frontier cohort is a proof-of-concept study in Flanders, Europe, that aims to support the development of a precision health-driven healthcare system, with a personalized approach in disease prevention. The study was designed to identify patterns, markers and processes that play a role in the spectrum between health and early onset of disease by combining information from a clinical setting, with multi-omics and continuous personal monitoring data.

Our preliminary findings in this cohort, which consists of healthy people with no disease diagnoses, show that physical disorders are still frequently present but not at a dysfunctional level (Suppl. figure 2) and that also the cardiovascular risk is, for example, increased in men (Suppl. table 1) (presumably due to increased BMI and cholesterol levels). Preliminary results indicate that self-reported physical symptoms don’t coincide to clinical events. This finding, and the combination of multiple omics datasets which contain more biological detail than the clinical measurements, will be further explored. Genetic analyses show the presence of pathogenic variants in the cohort that can benefit from early disease detection. Evaluation on how participants experienced these results will be reported later on.

Nonetheless, based on these results, it seems that other types of molecular entity parameters, like genomics, proteomics, metabolomics, etc., have significant added value to the currently established clinical data.

With regard to the longitudinal aspect, a noteworthy remark observed is the discomfort and inconvenience of a large sample donation at a clinical institute and the level of discomfort (e.g. venepuncture) that participating individuals experience. Although in our cohort we did not experience drop-outs as a result of this, in a larger trial, this is expected be a threshold for some to enter as well as affect the long-term viability of precision health cohorts. Therefore, efforts on multi-omics micro sampling methods that retain sample quality will be vital in the future. Very recently, a robust micro sampling approach was presented that allows individuals to collect small volumes of blood samples at home by themselves. As sample quality and parameter depth are not compromised, such an approach is a promising opportunity to improve sample collection and, thereby also, participant compliance in precision health cohort studies (10). Future research can implement this approach and fall back on our study with its unique longitudinal sampling frequency to assess what minimum (longitudinal) collection frequency and depth provide added value for early on-set health deviation detection.

Future research can implement this approach and fall back on our study with its unique longitudinal sampling frequency to assess what minimum (longitudinal) collection frequency and depth provide added value for early on-set health deviation detection.

The I Am Frontier is a POC cohort designed to explore what is needed in the long run to evolve towards a personalized prevention approach in a clinical setting (31). The various types of collected omics data form an ideal basis for that. In general, this study illustrates the viability of precision health in healthcare. However, more detailed analyses of this cohort, will be necessary to clarify which measurements are relevant for this purpose.

Due to privacy of the limited number of participants the data is only available for further research upon reasonable request. Requests will be processed on a study specific basis.

The privacy sensitive nature of the data did not allow it to be deposited into public repositories (32).

### Further details

#### Strengths and limitations

The I AM frontier cohort consists of deeply phenotyped patient samples with a profound sampling density (over 200 clinical parameters as well as omics technologies in blood, urine, stool, saliva as well as data collection through wearables, surveys and interviews) and unique longitudinal granularity. Therefore, the development of future precision health applications and the prevention of chronic diseases through prospective research can benefit from the availability of this biobank.

Whole genome sequencing is performed on participants, allowing for the creation of baseline risk profiles for genetic predisposition to common diseases. The study also includes extensive questionnaires on food intake, medical status, and perceptions of health from participants. Additionally, tracking data on physical activity is available for each participant. The questionnaire and tracking data can be integrated with clinical phenotype and genotype information to complement the molecular data analysis. Another important aspect is that participants’ perceptions of precision health implementations in healthcare, including ethical issues, are included in the scope of the cohort. Communication regarding the disclosure of the WGS test was a well-considered aspect of the cohort study. Guidelines or protocols for sharing this information with research participants are still in their early stages. For this POC, the research general practitioner (GP) took responsibility for conveying the results to the participants. However, when severely deviating results were found, the research GP first contacted the participant’s GP to discuss the findings. This protocol was evaluated before and after the cohort study in a WGS perception questionnaire.

One limitation of this study design was that data collection had to be stopped after 12 months, six months earlier than planned, due to the COVID-19 pandemic that arose in 2020, but this had no impact on the first year of data collection. We did not have any drop-outs in the first year. Only in the very last month of March 2020 ten drop-outs did occur. Via questionnaires, participants reported that our approach of giving feedback on certain parts of the results helped maintain their motivation to participate in the study. The cohort study consists of a limited number of 30 participants, so deep molecular profiling could be accomplished. This was done to facilitate efficient extrapolation for larger numbers of participants in future efforts. Since the implemented sampling technologies were selected to maximize overlap with large cross-sectional biobanks, the small sample size of the cohort can be compensated for by integrating with other cohort datasets, such as the UK Biobank. Such an integration allows comparison across different health and disease statuses (33). To optimally assess the long term potential of the data, integration of the I AM frontier cohort with several external datasets is being carried out at the time of writing.

## Data Availability

Due to participants' privacy, the data is available upon request to the I AM Frontier Project Data Access Committee for further research. The sensitive nature of the data does not allow it to be deposited into public repositories. We welcome collaboration with other research groups focused on personalized prevention, offering data sharing opportunities to advance research and improve healthcare outcomes.

## Declarations

### Ethics approval and consent to participate

The study was approved by the ethical committee of the Antwerp University Hospital (RegN°:B300201837314).

### Consent for publication

Not applicable

## Author contributions

Conceptualization: DH, NL, CDC, GB, JT, JH and GE; Formal analysis: DH, CDC, MP and ACR; Data Curation: AS, JH, GB; Writing – Original Draft: DH, NL, CDC, MP, GB, JT, GE; Writing – Review & Editing, DH, NL, CDC, RVDH, JH, GE, JT; Funding Acquisition & Supervision: NL, JH, GE; Project Administration: JT, GE, NL; Resources: GB, RVDH.

## Funding

This project was entirely funded by VITO NV.

## Acknowledgements

We want to thank:

Dr. Leo Geudens, our study doctor, for sharing his profound medical expertise and giving patients feedback. Hilde Bertels and Liesbeth Cayers for collecting samples, prof. Dr. Kristien Hens for sharing her extensive ethical knowledge, and Jordy Swysen for his tireless efforts in conducting participant interviews. Domus Medica advisory board for their valued feedback during the cohort period. Finally, Lotte Mollen, An Van Rompay, and Daniëlla Ooms for operationalizing the IAM Frontier cohort on a daily basis.

## List of abbreviations

ACMG: American College of Medical Genetics and Genomics
ADA: American diabetes association
BMI: Body mass index
CHIPS: Cohen-Hoberman Inventory of Physical Symptoms
CVD: Cardiovascular disease
EDTA: Ethylenediaminetetraacetic acid
GP: General practitioner
IPOP: Integrated personal omics profiling
MS: Mass spectrometry
NMR: Nuclear magnetic resonance
PBMC: Peripheral blood mononuclear cells
PCA: Principle component analyses
POC: Proof of concept
SNP: Single nucleotide polymorphism
WGS: Whole genome sequencing

## Notes

### Competing Interest Statement

The authors have declared no competing interest.

### Author Declarations

The study was approved by the ethical committee of the Antwerp University Hospital (RegNr:B300201837314).

