## Supplementary figures and images for "Cohort profile: The I AM Frontier prospective cohort study in Flanders"

### Supplementary figure 1

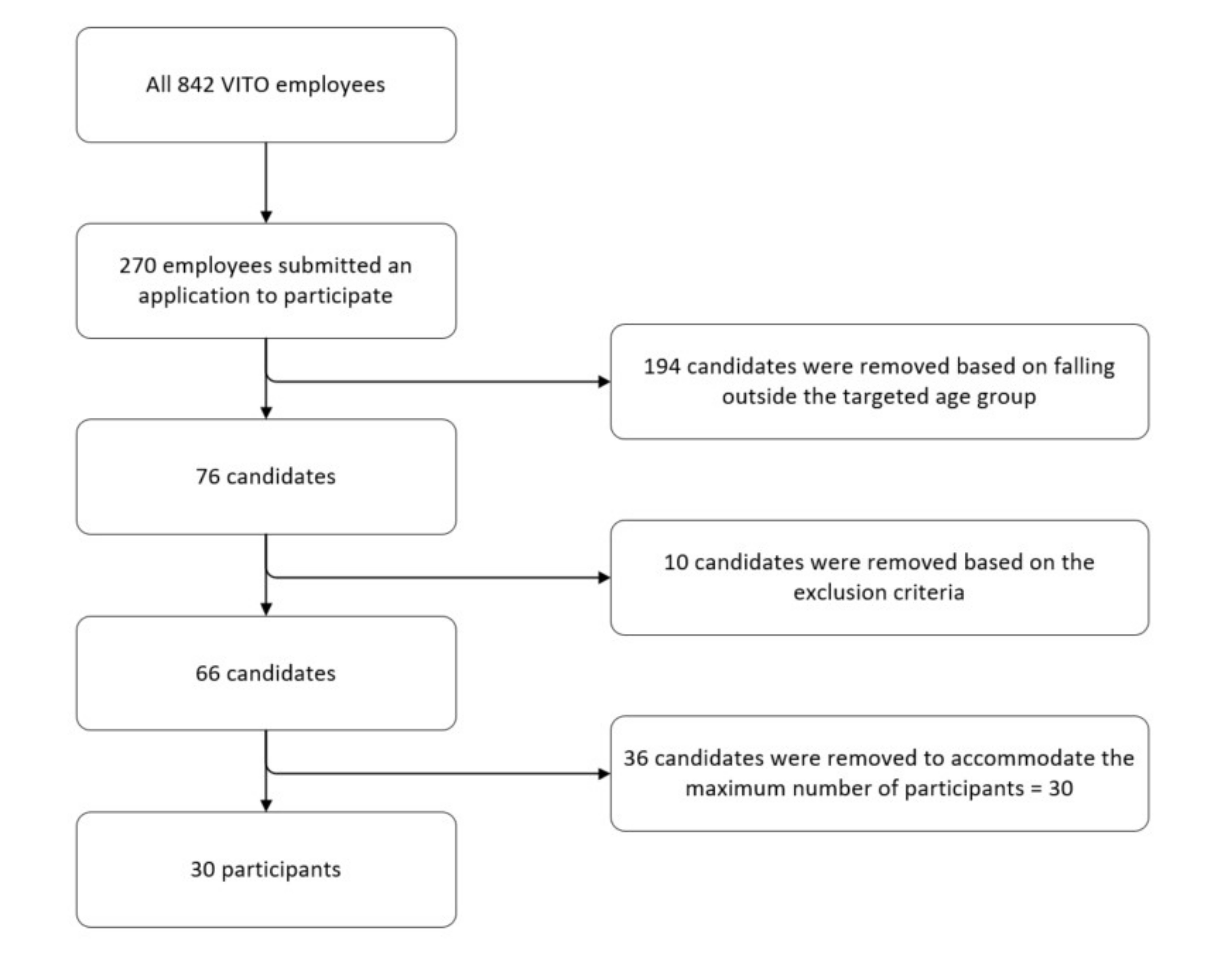

### Supplementary figure 2

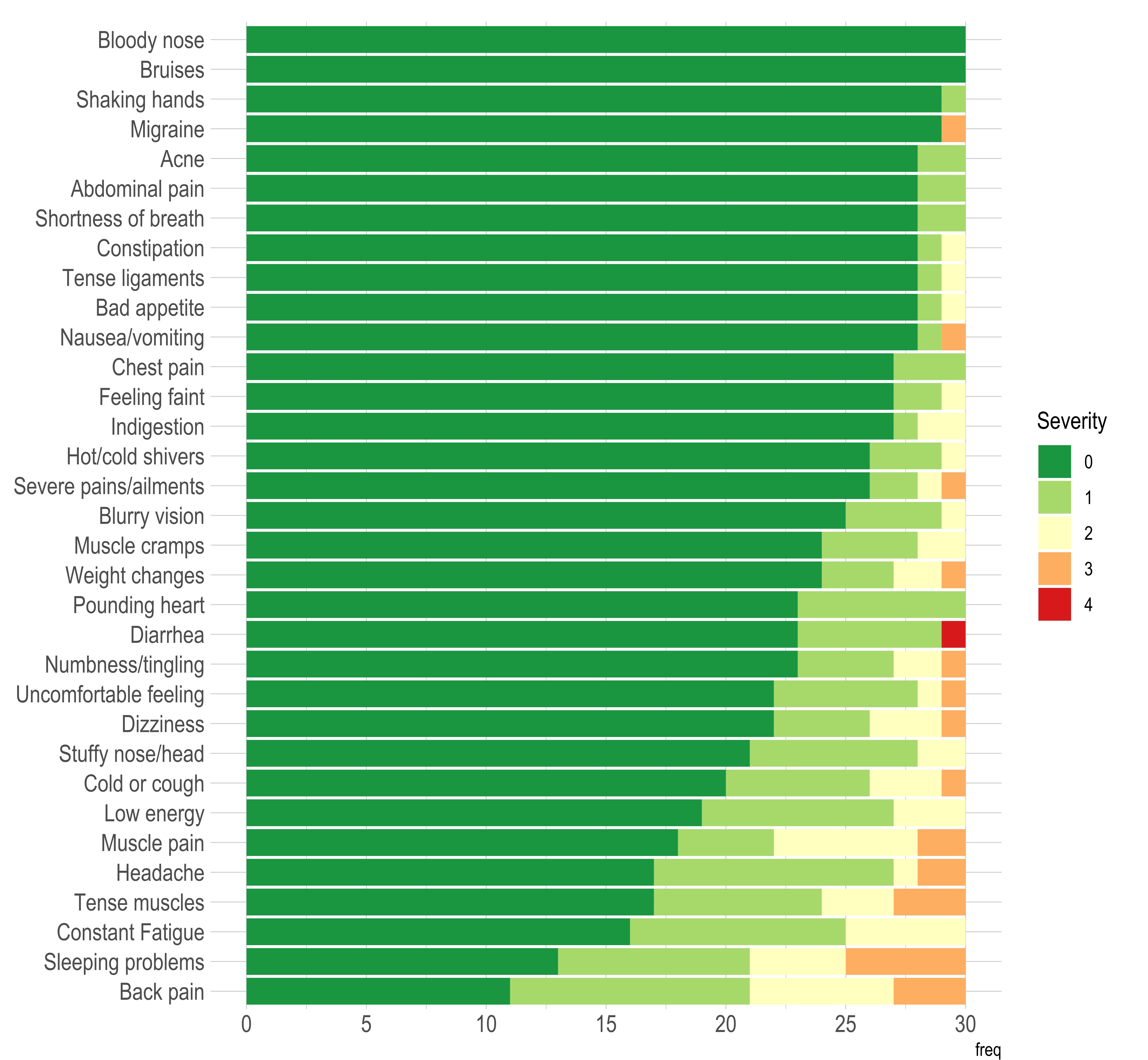

### Supplementary figure 3

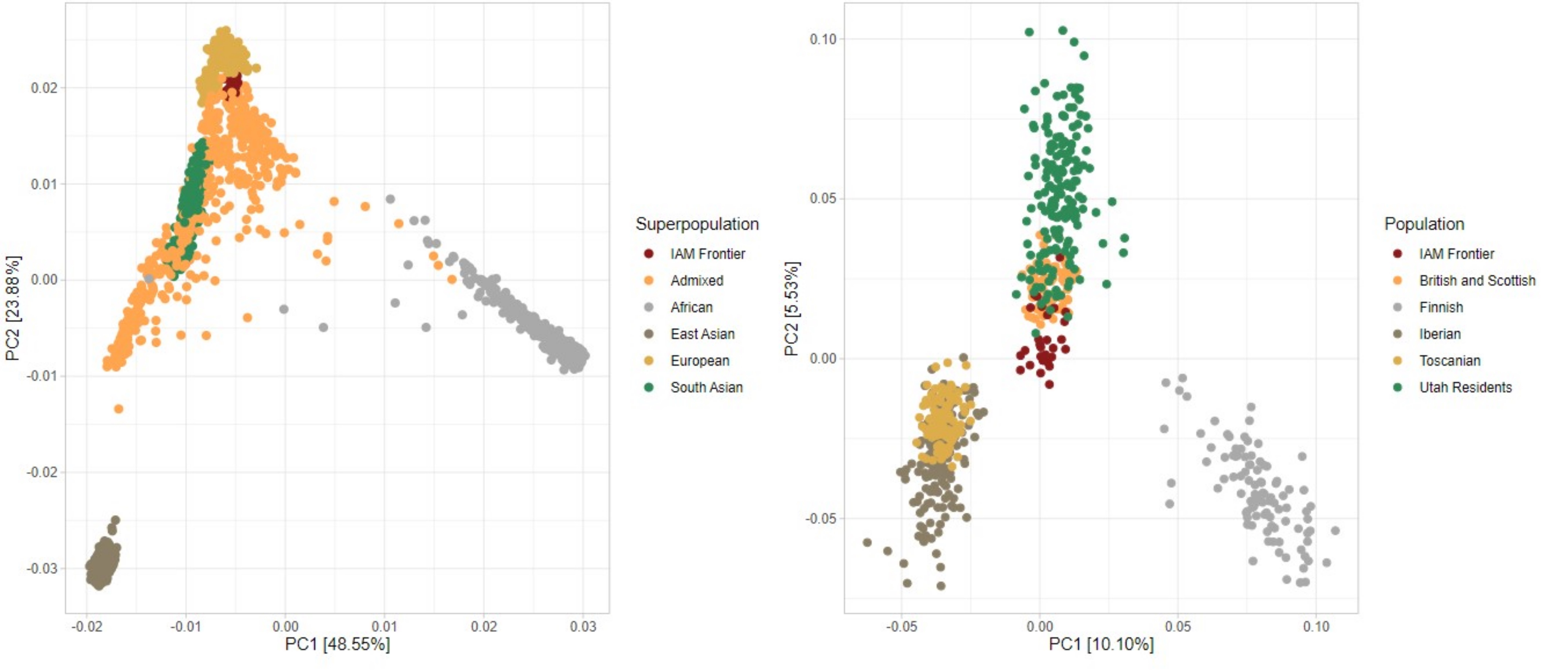

### Supplementary table 1

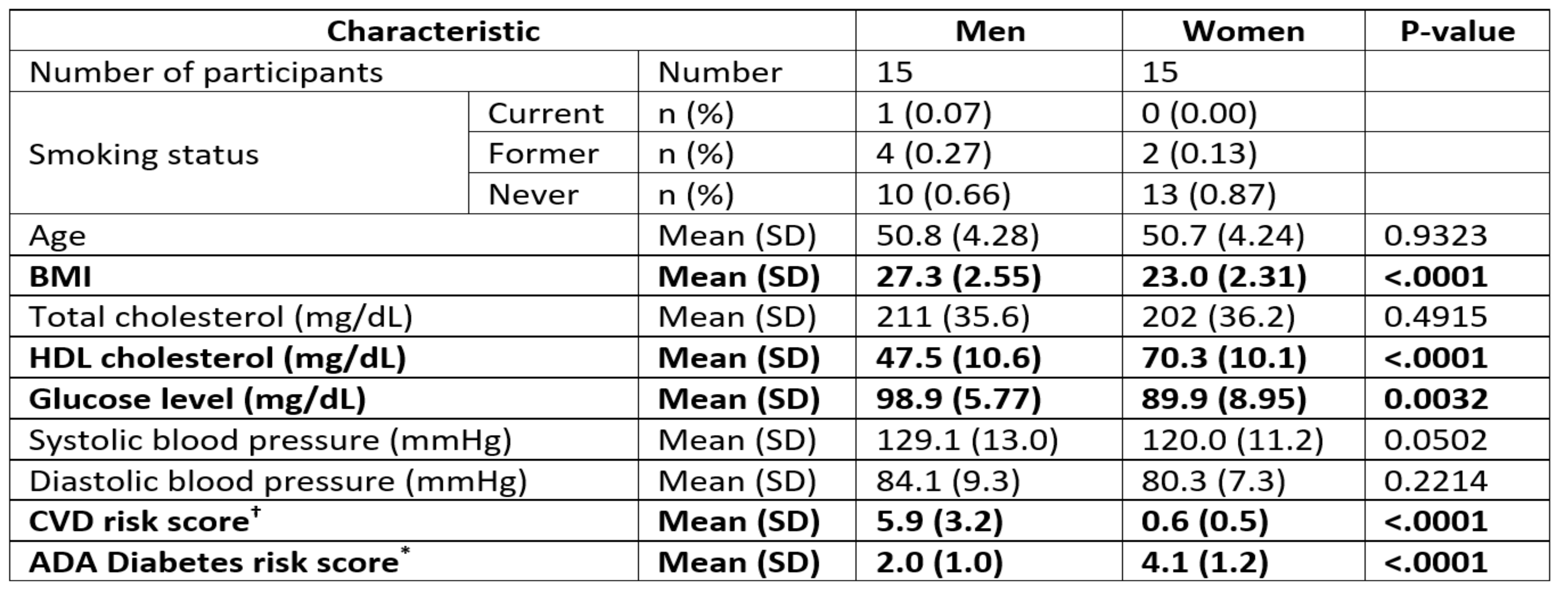
