## Supplementary table 2 for "Cohort profile: The I AM Frontier prospective cohort study in Flanders"

| Endpoint | Clinical parameters | | | | | | | | | | | | | Times tested |
|---|---|---|---|---|---|---|---|---|---|---|---|---|---|---|
| **Allergy** | gx3 Grasses | wx5 Herbs 1 | wx6 Herbs 2 | tx5 Trees 1 | tx6 Trees 2 | fx5 Food | | | | | | | | 1 |
| **Allergy** | mx1 Molds | ex1 Animal dander | d1 House dust mite | h1 House dust (Greer) | h2 House dust (Hollister) | f1 Egg white | f2 Cow's milk | f4 Wheat | f13 Peanut | f14 Soybean | f79 Gluten | c1 Penicilloyl G | c2 Penicilloyl V | 1 |
| **Allergy** | IgE | | | | | | | | | | | | | 8 |
| **Anemia** | Iron | Transferrin | Ferritin | Folic acid | Haptoglobin | | | | | | | | | 13 |
| **Antibodies directed against** | Adrenal gland | Skin | Insulin | Pancreas | Ac. Chol. Rec | Smooth muscle | LKM | Mitochondria | Intrinsic factor | Kidney (GBM) | | | | 1 |
| **Adrenal gland** | Transcortin | Cortisol | Aldosterone | Renin | | | | | | | | | | 13 |
| **Bleeding** | Factor VIII | Factor IX | APTT | Fibrinogen | von Willebrand factor activity | | Homocysteine | | | | | | | 1 |
| **Bone Metabolism** | Bone specific alkaline phosphatase | | Parathyroid hormone | | | | | | | | | | | 1 |
| **Bone Metabolism** | Vitamin D | | | | | | | | | | | | | 8 |
| **Cardiovascular** | Cholesterol | HDL Cholesterol | Triglycerides | LDL Cholesterol | Apolipoprotein A1/B | LP(a) | CK | Troponin | LDH | | | | | 13 |
| **Celiac disease** | Transglutaminase IgA | | | | | | | | | | | | | 1 |
| **Hormones** | IGF-I | PSA secreening/free PSA (M) | Progesterone/DHEA sulfate/Testosterone/SHBG (M+F) | | Estrone (F) | | | | | | | | | 1 |
| **Hormones** | Gastrin | | | | | | | | | | | | | 13 |
| **Immunohaematology & Immunology** | Blood type ABO-D E+S | Rhesus | Direct Antiglobulin | Irregular antibodies | Tryptase | | | | | | | | | 1 |
| **Immunology** | T, B, and NK cells | T cells, T4/T8 | | | | | | | | | | | | 8 |
| **Immunology** | IgG2 | IgG3 | IgG4 | Complement CH50 | Complement C3 | Complement C3d | Complement C4 | | | | | | | 13 |
| **Inflammation** | Sedimentation | | | | | | | | | | | | | 8 |
| **Inflammation** | Hemoglobin | Hematocrit | WBC | WBC formula | Platelets | Ultra sensitive CRP | | | | | | | | 13 |
| **Ionogram** | Sodium | Potassium | Chlorides | Bicarbonate | Calcium | Phosphate | Magnesium | Copper | Zinc | Osmolality | | | | 13 |
| **Carbohydrate metabolism** | Glucose (fasting) | HbA1c | Insuline (fasting) | C-peptide (fasting) | | | | | | | | | | 13 |
| **Liver** | Bilirubin total | AST | ALT | Gamma-GT | Alkaline phosphatase | LDH | Lipase | | | | | | | 13 |
| **Kidney** | Urea | Creatinine | Uric acid | | | | | | | | | | | 13 |
| **Orthomolecular tests** | Glutathione peroxidase | Total antioxidant status | | | | | | | | | | | | 1 |
| **Other** | Immunofixation | Calcitonin | ACTH | Angiotens. Conv. Enz. (ACE) | Cerulopasmin + copper | Cholinesterase | Transthyretin | Alph-1 antitrypsin | Beta-2 microglubulin | C1-inhibitor | BNP | TSI | | 1 |
| **Proteins** | Protein total | Protein electrophoresis | IgA | IgG | IGM | | | | | | | | | 13 |
| **Rheumatism - autoantibodies** | Rheumatoid factor (RF) | Anti-CCP | Antinuclear antibodies (ANA) | HLA B27 | | | | | | | | | | 1 |
| **Thyroid** | TSH | Free T4 | Anti-thyroglobulin | Thryoglobulin | | | | | | | | | | 8 |
| **Thrombosis** | D-Dimers | APC-resistance | Anti-cardiolipin IgM | Anti-cardiolipin IgG | Anti-B2 GPI IgM | Anti-B2 GPI IgG | | | | | | | | 1 |
| **Thrombosis** | Antithrombin | Protein C activity | Protein S free | | | | | | | | | | | 13 |
| **Vitamins** | Vitamin A | Vitamin C (**Month 4 and 10 only**) | | Vitamin E | Vitamin B12 | Vitamin B1 | Vitamin B2 | Vitamin B6 | B-Carotene | | | | | 8 |

| | | | | | | |
|---|---|---|---|---|---|---|
| *Color Code to distribute single time tests over time* | 1st month | 2nd month | 4th month | 6th month | 10th month | 12th month |
