## Supplementary table 3 for "Cohort profile: The I AM Frontier prospective cohort study in Flanders"

LB-0428 11/2019

### Nightingale Blood Biomarker Analysis Service

*Boost your research with comprehensive blood metabolite insights.*

#### Powerful platform enabled by NMR

##### Unique combination of biomarkers

Biomarkers include clinically validated routine markers, emerging biomarkers with strong medical relevance and promising, new biomarkers.

##### Robust and highly reproducible results

Our fully automated analysis process is constantly monitored under a certified quality management system. And the NMR technology allows high reproducibility which ensures consistent and reliable results across all sample sets.

##### Fast, cost-efficient and scalable technology

We use a high-throughput NMR technology which ensures efficient analysis for sample sets of all sizes without batch effects.

##### Accurate and fully quantified metabolic data

Not only our analysis, but even our quantification process of the NMR spectral data is fully automated, which provides precise and accurate metabolite results in absolute concentration units.

##### Comprehensive overview of an individual's health

Biomarkers in our panel provide a physiologically meaningful picture of the overall health making it possible to explore novel connections between metabolites and an individual's health status.

##### Certified quality management system

Nightingale's quality management system has been certified according to EN ISO 13485 standard.

#### Tech specifications

| | |
|---|---|
| **Technology** | $^1$H NMR Spectroscopy, Nightingale Health's proprietary analysis |
| **Specimen type** | Serum and Plasma |
| **Sample volume** | 100 µL and 350 µL |
| **Number of biomarkers** | 250 |
| **Result units** | Absolute biomarker quantification (mmol/l or g/l) |
| **Sample storage** | Long-term storage -70˚C or below |

#### Clinically validated biomarkers

| | |
|---|---|
| **Cholesterol** | |
| Total cholesterol | mmol/l |
| VLDL cholesterol | mmol/l |
| Clinical LDL cholesterol | mmol/l |
| HDL cholesterol | mmol/l |
| **Triglycerides** | |
| Total triglycerides | mmol/l |
| **Apolipoproteins** | |
| Apolipoprotein B | g/l |
| Apolipoprotein A1 | g/l |
| Ratio of apolipoprotein B to apolipoprotein A1 | ratio |
| **Fatty acids** | |
| Total fatty acids | mmol/l |
| Omega-3 fatty acids | mmol/l |
| Omega-6 fatty acids | mmol/l |
| Monounsaturated fatty acids | mmol/l |
| Saturated fatty acids | mmol/l |
| **Fatty acid ratios** | |
| Ratio of omega-3 fatty acids to total fatty acids | % |
| Ratio of omega-6 fatty acids to total fatty acids | % |
| Ratio of monounsaturated fatty acids to total fatty acids | % |
| Ratio of saturated fatty acids to total fatty acids | % |
| **Branched-chain amino acids** | |
| Isoleucine | mmol/l |
| Leucine | mmol/l |
| Valine | mmol/l |
| **Glycolysis related metabolites** | |
| Glucose | mmol/l |
| **Fluid balance** | |
| Creatinine | mmol/l |
| **Inflammation** | |
| Glycoprotein acetyls | mmol/l |

#### List of all biomarkers

| Metabolite | Unit |
|---|---|
| **Cholesterol** | |
| Total cholesterol | mmol/l |
| Total cholesterol minus HDL-C | mmol/l |
| Remnant cholesterol (non-HDL, non-LDL -cholesterol) | mmol/l |
| VLDL cholesterol | mmol/l |
| Clinical LDL cholesterol | mmol/l |
| LDL cholesterol | mmol/l |
| HDL cholesterol | mmol/l |
| **Triglycerides** | |
| Total triglycerides | mmol/l |
| Triglycerides in VLDL | mmol/l |
| Triglycerides in LDL | mmol/l |
| Triglycerides in HDL | mmol/l |
| **Phospholipids** | |
| Total phospholipids in lipoprotein particles | mmol/l |
| Phospholipids in VLDL | mmol/l |
| Phospholipids in LDL | mmol/l |
| Phospholipids in HDL | mmol/l |
| **Cholesteryl esters** | |
| Total esterified cholesterol | mmol/l |
| Cholesterol esters in VLDL | mmol/l |
| Cholesterol esters in LDL | mmol/l |
| Cholesterol esters in HDL | mmol/l |
| **Free cholesterol** | |
| Total free cholesterol | mmol/l |
| Free cholesterol in VLDL | mmol/l |
| Free cholesterol in LDL | mmol/l |
| Free cholesterol in HDL | mmol/l |
| **Total lipids** | |
| Total lipids in lipoprotein particles | mmol/l |
| Total lipids in VLDL | mmol/l |
| Total lipids in LDL | mmol/l |
| Total lipids in HDL | mmol/l |
| **Lipoprotein particle concentrations** | |
| Total concentration of lipoprotein particles | mmol/l |
| Concentration of VLDL particles | mmol/l |
| Concentration of LDL particles | mmol/l |
| Concentration of HDL particles | mmol/l |
| **Lipoprotein particle sizes** | |
| Average diameter for VLDL particles | nm |
| Average diameter for LDL particles | nm |
| Average diameter for HDL particles | nm |

| Metabolite | Unit |
|---|---|
| **Other lipids** | |
| Phosphoglycerides | mmol/l |
| Ratio of triglycerides to phosphoglycerides | ratio |
| Total cholines | mmol/l |
| Phosphatidylcholines | mmol/l |
| Sphingomyelins | mmol/l |
| **Apolipoproteins** | |
| Apolipoprotein B | g/l |
| Apolipoprotein A1 | g/l |
| Ratio of apolipoprotein B to apolipoprotein A1 | ratio |
| **Fatty acids** | |
| Total fatty acids | mmol/l |
| Degree of unsaturation | degree |
| Omega-3 fatty acids | mmol/l |
| Omega-6 fatty acids | mmol/l |
| Polyunsaturated fatty acids | mmol/l |
| Monounsaturated fatty acids | mmol/l |
| Saturated fatty acids | mmol/l |
| Linoleic acid | mmol/l |
| Docosahexaenoic acid | mmol/l |
| **Fatty acid ratios** | |
| Ratio of omega-3 fatty acids to total fatty acids | % |
| Ratio of omega-6 fatty acids to total fatty acids | % |
| Ratio of polyunsaturated fatty acids to total fatty acids | % |
| Ratio of monounsaturated fatty acids to total fatty acids | % |
| Ratio of saturated fatty acids to total fatty acids | % |
| Ratio of linoleic acid to total fatty acids | % |
| Ratio of docosahexaenoic acid to total fatty acids | % |
| Ratio of polyunsaturated fatty acids to monounsaturated fatty acids | ratio |
| Ratio of omega-6 fatty acids to omega-3 fatty acids | ratio |
| **Amino acids** | |
| Alanine | mmol/l |
| Glutamine | mmol/l |
| Glycine | mmol/l |
| Histidine | mmol/l |
| **Branched-chain amino acids** | |
| Total concentration of branded -chain amino acids (leucine + isoleucine + valine) | mmol/l |
| Isoleucine | mmol/l |
| Leucine | mmol/l |
| Valine | mmol/l |
| **Aromatic amino acids** | |
| Phenylalanine | mmol/l |
| Tyrosine | mmol/l |

| Metabolite | Unit |
|---|---|
| **Glycolysis related metabolites** | |
| Glucose | mmol/l |
| Lactate | mmol/l |
| Pyruvate | mmol/l |
| Citrate ** | mmol/l |
| Glycerol * | mmol/l |
| **Ketone bodies** | |
| 3-Hydroxybutyrate | mmol/l |
| Acetate | mmol/l |
| Acetoacetate | mmol/l |
| Acetone | mmol/l |
| **Fluid balance** | |
| Creatinine | mmol/l |
| Albumin | g/l |
| **Inflammation** | |
| Glycoprotein acetyls | mmol/l |
| **Lipoprotein subclasses** | |
| **Chylomicrons and extremely large VLDL (particle diameters from 75 nm upwards)** | |
| Concentration of chylomicrons and extremely large VLDL particles | mmol/l |
| Total lipids in chylomicrons and extremely large VLDL | mmol/l |
| Phospholipids in chylomicrons and extremely large VLDL | mmol/l |
| Cholesterol in chylomicrons and extremely large VLDL | mmol/l |
| Cholesteryl esters in chylomicrons and extremely large VLDL | mmol/l |
| Free cholesterol in chylomicrons and extremely large VLDL | mmol/l |
| Triglycerides in chylomicrons and extremely large VLDL | mmol/l |
| **Very large VLDL (average diameter 64 nm)** | |
| Concentration of very large VLDL particles | mmol/l |
| Total lipids in very large VLDL | mmol/l |
| Phospholipids in very large VLDL | mmol/l |
| Cholesterol in very large VLDL | mmol/l |
| Cholesteryl esters in very large VLDL | mmol/l |
| Free cholesterol in very large VLDL | mmol/l |
| Triglycerides in very large VLDL | mmol/l |
| **Large VLDL (average diameter 53.6 nm)** | |
| Concentration of large VLDL particles | mmol/l |
| Total lipids in large VLDL | mmol/l |
| Phospholipids in large VLDL | mmol/l |
| Cholesterol in large VLDL | mmol/l |
| Cholesteryl esters in large VLDL | mmol/l |
| Free cholesterol in large VLDL | mmol/l |
| Triglycerides in large VLDL | mmol/l |

| Metabolite | Unit |
|---|---|
| **Medium VLDL (average diameter 44.5 nm)** | |
| Concentration of medium VLDL particles | mmol/l |
| Total lipids in medium VLDL | mmol/l |
| Phospholipids in medium VLDL | mmol/l |
| Cholesterol in medium VLDL | mmol/l |
| Cholesteryl esters in medium VLDL | mmol/l |
| Free cholesterol in medium VLDL | mmol/l |
| Triglycerides in medium VLDL | mmol/l |
| **Small VLDL (average diameter 36.8 nm)** | |
| Concentration of small VLDL particles | mmol/l |
| Total lipids in small VLDL | mmol/l |
| Phospholipids in small VLDL | mmol/l |
| Cholesterol in small VLDL | mmol/l |
| Cholesteryl esters in small VLDL | mmol/l |
| Free cholesterol in small VLDL | mmol/l |
| Triglycerides in small VLDL | mmol/l |
| **Very small VLDL (average diameter 31.3 nm)** | |
| Concentration of very small VLDL particles | mmol/l |
| Total lipids in very small VLDL | mmol/l |
| Phospholipids in very small VLDL | mmol/l |
| Cholesterol in very small VLDL | mmol/l |
| Cholesteryl esters in very small VLDL | mmol/l |
| Free cholesterol in very small VLDL | mmol/l |
| Triglycerides in very small VLDL | mmol/l |
| **IDL (average diameter 28.6 nm)** | |
| Concentration of IDL particles | mmol/l |
| Total lipids in IDL | mmol/l |
| Phospholipids in IDL | mmol/l |
| Cholesterol in IDL | mmol/l |
| Cholesteryl esters in IDL | mmol/l |
| Free cholesterol in IDL | mmol/l |
| Triglycerides in IDL | mmol/l |
| **Large LDL (average diameter 25.5 nm)** | |
| Concentration of large LDL particles | mmol/l |
| Total lipids in large LDL | mmol/l |
| Phospholipids in large LDL | mmol/l |
| Cholesterol in large LDL | mmol/l |
| Cholesteryl esters in large LDL | mmol/l |
| Free cholesterol in large LDL | mmol/l |
| Triglycerides in large LDL | mmol/l |
| **Medium LDL (average diameter 23 nm)** | |
| Concentration of medium LDL particles | mmol/l |
| Total lipids in medium LDL | mmol/l |
| Phospholipids in medium LDL | mmol/l |
| Cholesterol in medium LDL | mmol/l |
| Cholesteryl esters in medium LDL | mmol/l |
| Free cholesterol in medium LDL | mmol/l |
| Triglycerides in medium LDL | mmol/l |

*All listed biomarkers are available for Serum and Heparin plasma samples.*

*Biomarkers marked with * are not available for EDTA plasma samples.*
*Biomarkers marked with ** are not available for Citrate plasma samples.*

### List of Biomarkers

| Metabolite | Unit |
|---|---|
| **Small LDL (average diameter 18.7 nm)** | |
| Concentration of small LDL particles | mmol/l |
| Total lipids in small LDL | mmol/l |
| Phospholipids in small LDL | mmol/l |
| Cholesterol in small LDL | mmol/l |
| Cholesteryl esters in small LDL | mmol/l |
| Free cholesterol in small LDL | mmol/l |
| Triglycerides in small LDL | mmol/l |
| **Very large HDL (average diameter 14.3 nm)** | |
| Concentration of very large HDL particles | mmol/l |
| Total lipids in very large HDL | mmol/l |
| Phospholipids in very large HDL | mmol/l |
| Cholesterol in very large HDL | mmol/l |
| Cholesteryl esters in very large HDL | mmol/l |
| Free cholesterol in very large HDL | mmol/l |
| Triglycerides in very large HDL | mmol/l |
| **Large HDL (average diameter 12.1 nm)** | |
| Concentration of large HDL particles | mmol/l |
| Total lipids in large HDL | mmol/l |
| Phospholipids in large HDL | mmol/l |
| Cholesterol in large HDL | mmol/l |
| Cholesteryl esters in large HDL | mmol/l |
| Free cholesterol in large HDL | mmol/l |
| Triglycerides in large HDL | mmol/l |
| **Medium HDL (average diameter 10.9 nm)** | |
| Concentration of medium HDL particles | mmol/l |
| Total lipids in medium HDL | mmol/l |
| Phospholipids in medium HDL | mmol/l |
| Cholesterol in medium HDL | mmol/l |
| Cholesteryl esters in medium HDL | mmol/l |
| Free cholesterol in medium HDL | mmol/l |
| Triglycerides in medium HDL | mmol/l |
| **Small HDL (average diameter 8.7 nm)** | |
| Concentration of small HDL particles | mmol/l |
| Total lipids in small HDL | mmol/l |
| Phospholipids in small HDL | mmol/l |
| Cholesterol in small HDL | mmol/l |
| Cholesteryl esters in small HDL | mmol/l |
| Free cholesterol in small HDL | mmol/l |
| Triglycerides in small HDL | mmol/l |
| **Relative lipoprotein lipid concentrations** | |
| **Chylomicrons and extremely large VLDL ratios** | |
| Phospholipids to total lipids ratio in chylomicrons and extremely large VLDL | % |
| Cholesterol to total lipids ratio in chylomicrons and extremely large VLDL | % |
| Cholesteryl esters to total lipids ratio in chylomicrons and extremely large VLDL | % |
| Free cholesterol to total lipids ratio in chylomicrons and extremely large VLDL | % |
| Triglycerides to total lipids ratio in chylomicrons and extremely large VLDL | % |
| **Very large VLDL ratios** | |
| Phospholipids to total lipids ratio in very large VLDL | % |
| Cholesterol to total lipids ratio in very large VLDL | % |
| Cholesteryl esters to total lipids ratio in very large VLDL | % |
| Free cholesterol to total lipids ratio in very large VLDL | % |
| Triglycerides to total lipids ratio in very large VLDL | % |
| **Large VLDL ratios** | |
| Phospholipids to total lipids ratio in large VLDL | % |
| Cholesterol to total lipids ratio in large VLDL | % |
| Cholesteryl esters to total lipids ratio in large VLDL | % |
| Free cholesterol to total lipids ratio in large VLDL | % |
| Triglycerides to total lipids ratio in large VLDL | % |
| **Medium VLDL ratios** | |
| Phospholipids to total lipids ratio in medium VLDL | % |
| Cholesterol to total lipids ratio in medium VLDL | % |
| Cholesteryl esters to total lipids ratio in medium VLDL | % |
| Free cholesterol to total lipids ratio in medium VLDL | % |
| Triglycerides to total lipids ratio in medium VLDL | % |
| **Small VLDL ratios** | |
| Phospholipids to total lipids ratio in small VLDL | % |
| Cholesterol to total lipids ratio in small VLDL | % |
| Cholesteryl esters to total lipids ratio in small VLDL | % |
| Free cholesterol to total lipids ratio in small VLDL | % |
| Triglycerides to total lipids ratio in small VLDL | % |
| **Very small VLDL ratios** | |
| Phospholipids to total lipids ratio in very small VLDL | % |
| Cholesterol to total lipids ratio in very small VLDL | % |
| Cholesteryl esters to total lipids ratio in very small VLDL | % |
| Free cholesterol to total lipids ratio in very small VLDL | % |
| Triglycerides to total lipids ratio in very small VLDL | % |
| **IDL ratios** | |
| Phospholipids to total lipids ratio in IDL | % |
| Cholesterol to total lipids ratio in IDL | % |
| Cholesteryl esters to total lipids ratio in IDL | % |
| Free cholesterol to total lipids ratio in IDL | % |
| Triglycerides to total lipids ratio in IDL | % |
| **Large LDL ratios** | |
| Phospholipids to total lipids ratio in large LDL | % |
| Cholesterol to total lipids ratio in large LDL | % |
| Cholesteryl esters to total lipids ratio in large LDL | % |
| Free cholesterol to total lipids ratio in large LDL | % |
| Triglycerides to total lipids ratio in large LDL | % |
| **Medium LDL ratios** | |
| Phospholipids to total lipids ratio in medium LDL | % |
| Cholesterol to total lipids ratio in medium LDL | % |
| Cholesteryl esters to total lipids ratio in medium LDL | % |
| Free cholesterol to total lipids ratio in medium LDL | % |
| Triglycerides to total lipids ratio in medium LDL | % |
| **Small LDL ratios** | |
| Phospholipids to total lipids ratio in small LDL | % |
| Cholesterol to total lipids ratio in small LDL | % |
| Cholesteryl esters to total lipids ratio in small LDL | % |
| Free cholesterol to total lipids ratio in small LDL | % |
| Triglycerides to total lipids ratio in small LDL | % |
| **Very large HDL ratios** | |
| Phospholipids to total lipids ratio in very large HDL | % |
| Cholesterol to total lipids ratio in very large HDL | % |
| Cholesteryl esters to total lipids ratio in very large HDL | % |
| Free cholesterol to total lipids ratio in very large HDL | % |
| Triglycerides to total lipids ratio in very large HDL | % |
| **Large HDL ratios** | |
| Phospholipids to total lipids ratio in large HDL | % |
| Cholesterol to total lipids ratio in large HDL | % |
| Cholesteryl esters to total lipids ratio in large HDL | % |
| Free cholesterol to total lipids ratio in large HDL | % |
| Triglycerides to total lipids ratio in large HDL | % |
| **Medium HDL ratios** | |
| Phospholipids to total lipids ratio in medium HDL | % |
| Cholesterol to total lipids ratio in medium HDL | % |
| Cholesteryl esters to total lipids ratio in medium HDL | % |
| Free cholesterol to total lipids ratio in medium HDL | % |
| Triglycerides to total lipids ratio in medium HDL | % |
| **Small HDL ratios** | |
| Phospholipids to total lipids ratio in small HDL | % |
| Cholesterol to total lipids ratio in small HDL | % |
| Cholesteryl esters to total lipids ratio in small HDL | % |
| Free cholesterol to total lipids ratio in small HDL | % |
| Triglycerides to total lipids ratio in small HDL | % |

*All listed biomarkers are available for Serum and Heparin plasma samples.*

*Biomarkers marked with * are not available for EDTA plasma samples.*
*Biomarkers marked with ** are not available for Citrate plasma samples.*

Nightingale

Nightingale Health Ltd. provides a NMR (Nuclear Magnetic Resonance) based metabolomics technology, supplying biomarker analysis services for human blood, urine, CSF and umbilical cord blood samples. By measuring biomarkers from multiple pathways in a single experiment, Nightingale equips public health researchers with comprehensive insights into the effects of lifestyle factors and future disease risk, accelerating future breakthroughs in precision medicine. In the long term, the company plans to fully integrate its services into clinical practice, helping to empower patients to follow-up on their own well-being and take proactive steps to stay healthy.

#### See also


Nightingale CSF Biomarker Analysis Service
Nightingale Urine Biomarker Analysis Service
Nightingale Cord Blood Biomarker Analysis Service

LB-0428 11/2019

### Nightingale Urine Biomarker Analysis Service

*Fully quantitative metabolic data for novel scientific discoveries powered by recent breakthrough in metabolomics technology*

#### Powerful platform enabled by NMR

##### Robust and highly reproducible results

Our fully automated analysis process is constantly monitored. NMR technology allows high reproducibility which ensures consistent and reliable results across all sample sets.

##### Fast, cost-efficient and scalable technology

We use a high-throughput NMR technology which ensures efficient analysis for sample sets of all sizes without batch effects.

##### Accurate and fully quantified metabolic data

Not only our measurement of the samples, but also our quantification process of the NMR spectral data is fully automated, which provides precise and accurate metabolite results in absolute concentration units.

##### Comprehensive overview of an individual's health

Biomarkers in our panel provide a physiologically meaningful picture of the overall health making it possible to explore novel connections between metabolites and an individual's health status.

#### Tech specifications

| | |
|---|---|
| **Technology/ method** | 1H NMR Spectroscopy, Nightingale Health's proprietary analysis |
| **Sample volume** | 500 µL |
| **Number of biomarkers** | 54 |
| **Result units** | Absolute biomarker quantification (mmol/l and ratio to creatinine) |
| **Required sample storage** | Long-term storage -70˚C or below |

#### Application examples

- Early risk detection and prognostics of type 1 and type 2 diabetes as well as diabetic complications, especially for diabetic kidney disease.
- Molecular understanding of cardiometabolic risk factors such as adiposity and body fat distribution, and what role they play in the disease etiology.
- Genetic regulation of urine metabolism and further implications to disease etiology
- Exploring metabolic effects of an individual's diet and lifestyle on health

#### List of Biomarkers

Available in March 2020

| Metabolite | Unit |
|---|---|
| **Amino acids** | |
| Alanine | mmol/l & ratio to creatinine |
| Glutamine | mmol/l & ratio to creatinine |
| Glycine | mmol/l & ratio to creatinine |
| Histidine | mmol/l & ratio to creatinine |
| Taurine | mmol/l & ratio to creatinine |
| Threonine | mmol/l & ratio to creatinine |
| Tryptophan | mmol/l & ratio to creatinine |
| *Branched-chain amino acids* | |
| Isoleucine | mmol/l & ratio to creatinine |
| Leucine | mmol/l & ratio to creatinine |
| Valine | mmol/l & ratio to creatinine |
| *Aromatic amino acids* | |
| Tyrosine | mmol/l & ratio to creatinine |
| **Dietary metabolites** | |
| 2-Furoylglycine | mmol/l & ratio to creatinine |
| 3-Methylhistidine | mmol/l & ratio to creatinine |
| Arabinose | mmol/l & ratio to creatinine |
| Ethanol | mmol/l & ratio to creatinine |
| HPHPA | mmol/l & ratio to creatinine |
| Mannitol | mmol/l & ratio to creatinine |
| Proline betaine* | mmol/l & ratio to creatinine |
| Propylene glycol* | mmol/l & ratio to creatinine |
| Quinic acid | mmol/l & ratio to creatinine |
| Sucrose | mmol/l & ratio to creatinine |
| Trans-aconitate | mmol/l & ratio to creatinine |
| Xanthosine | mmol/l & ratio to creatinine |
| Xylose | mmol/l & ratio to creatinine |

| Metabolite | Unit |
|---|---|
| **Fluid balance** | |
| Creatinine | mmol/l |
| **Glycolysis related metabolites** | |
| cis-Aconitate | mmol/l & ratio to creatinine |
| Citrate | mmol/l & ratio to creatinine |
| Glucose | mmol/l & ratio to creatinine |
| Lactate | mmol/l & ratio to creatinine |
| **Ketone bodies** | |
| Acetate | mmol/l & ratio to creatinine |
| **Microbial metabolism** | |
| 3-Hydroxyhippurate | mmol/l & ratio to creatinine |
| Dimethylamine | mmol/l & ratio to creatinine |
| Trimethylamine N-oxide | mmol/l & ratio to creatinine |
| **Miscellaneous** | |
| 2-Hydroxyisobutyrate | mmol/l & ratio to creatinine |
| 3-Hydroxyisobutyrate | mmol/l & ratio to creatinine |
| 3-Hydroxyisovalerate | mmol/l & ratio to creatinine |
| 4-Deoxyerythronic acid | mmol/l & ratio to creatinine |
| 4-Deoxythreonate | mmol/l & ratio to creatinine |
| 4-Hydroxyhippurate | mmol/l & ratio to creatinine |
| Allantoin | mmol/l & ratio to creatinine |
| Creatine | mmol/l & ratio to creatinine |
| Ethanolamine | mmol/l & ratio to creatinine |
| Formate | mmol/l & ratio to creatinine |
| Glycolate | mmol/l & ratio to creatinine |
| Hypoxanthine | mmol/l & ratio to creatinine |
| Indoxyl Sulfate | mmol/l & ratio to creatinine |
| Pseudouridine | mmol/l & ratio to creatinine |
| Pyroglutamate | mmol/l & ratio to creatinine |
| Urea | mmol/l & ratio to creatinine |

| Metabolite | Unit |
|---|---|
| **Nicotinate and nicotinamide metabolism** | |
| 1-Methylnicotinamide | mmol/l & ratio to creatinine |
| Trigonelline | mmol/l & ratio to creatinine |
| **Phenylalanine metabolism** | |
| Hippurate | mmol/l & ratio to creatinine |
| **Pyrimidine metabolism** | |
| 3-Aminoisobutyrate | mmol/l & ratio to creatinine |
| Uracil | mmol/l & ratio to creatinine |

** Preliminary biomarkers*

Nightingale

Nightingale Health Ltd. provides a NMR (Nuclear Magnetic Resonance) based metabolomics technology, supplying biomarker analysis services for human blood, urine, CSF and umbilical cord blood samples. By measuring biomarkers from multiple pathways in a single experiment, Nightingale equips public health researchers with comprehensive insights into the effects of lifestyle factors and future disease risk, accelerating future breakthroughs in precision medicine. In the long term, the company plans to fully integrate its services into clinical practice, helping to empower patients to follow-up on their own well-being and take proactive steps to stay healthy.

##### See also


Nightingale CSF Biomarker Analysis Service
Nightingale Blood Analysis Service
Nightingale Cord Blood Biomarker Analysis Service
