## Supplementary table 4 for "Cohort profile: The I AM Frontier prospective cohort study in Flanders"

Olink®

Protein assay list

### Olink® Target 96 Cardiometabolic

Product number: 95360

| | | | |
|---|---|---|---|
| Angiogenin (ANG) | P03950 | Endoglin (ENG) | P17813 |
| Angiopoietin-related protein 3 (ANGPTL3) | Q9Y5C1 | Fetuin-B (FETUB) | Q9UGM5 |
| Apolipoprotein M (APOM) | O95445 | Ficolin-2 (FCN2) | Q15485 |
| Beta-Ala-His dipeptidase (CNDP1) | Q96KN2 | Glutaminyl-peptide cyclotransferase (QPCT) | Q16769 |
| Beta-galactoside alpha-2,6-sialyltransferase 1 (ST6GAL1) | P15907 | Granulysin (GNLY) | P22749 |
| Cadherin-1 (CDH1) | P12830 | Growth arrest-specific protein 6 (GAS6) | Q14393 |
| Carbonic anhydrase 1 (CA1) | P00915 | Hepatocyte growth factor receptor (MET) | P08581 |
| Carbonic anhydrase 3 (CA3) | P07451 | Ig lambda-2 chain C regions (IGLC2) | P0CG05 |
| Carbonic anhydrase 4 (CA4) | P22748 | Insulin-like growth factor-binding protein 3 (IGFBP3) | P17936 |
| Cartilage acidic protein 1 (CRTAC1) | Q9NQ79 | Insulin-like growth factor-binding protein 6 (IGFBP6) | P24592 |
| Cartilage oligomeric matrix protein (COMP) | P49747 | Integrin alpha-M (ITGAM) | P11215 |
| C-C motif chemokine 5 (CCL5) | P13501 | Intercellular adhesion molecule 1 (ICAM1) | P05362 |
| C-C motif chemokine 14 (CCL14) | Q16627 | Intercellular adhesion molecule 3 (ICAM3) | P32942 |
| C-C motif chemokine 18 (CCL18) | P55774 | Interleukin-7 receptor subunit alpha (IL7R) | P16871 |
| CD59 glycoprotein (CD59) | P13987 | Latent-transforming growth factor beta-binding protein 2 (LTBP2) | Q14767 |
| Coagulation factor VII (F7) | P08709 | Leukocyte immunoglobulin-like receptor subfamily B member 1 (LILRB1) | Q8NHL6 |
| Coagulation factor XI (F11) | P03951 | Leukocyte immunoglobulin-like receptor subfamily B member 2 (LILRB2) | Q8N423 |
| Collagen alpha-1(XVIII) chain (COL18A1) | P39060 | Leukocyte immunoglobulin-like receptor subfamily B member 5 (LILRB5) | O75023 |
| Complement C1q tumor necrosis factor-related protein 1 (C1QTNF1) | Q9BXJ1 | Lithostathine-1-alpha (REG1A) | P05451 |
| Complement C2 (C2) | P06681 | Liver carboxylesterase 1 (CES1) | P23141 |
| Complement factor H-related protein 5 (CFHR5) | Q9BXR6 | Low affinity immunoglobulin gamma Fc region receptor II-a (FCGR2A) | P12318 |
| Complement receptor type 2 (CR2) | P20023 | Low affinity immunoglobulin gamma Fc region receptor III-B (FCGR3B) | O75015 |
| Cystatin-C (CST3) | P01034 | L-selectin (SELL) | P14151 |
| Dipeptidyl peptidase 4 (DPP4) | P27487 | | |
| EGF-containing fibulin-like extracellular matrix protein 1 (EFEMP1) | Q12805 | | |

Table continues on reverse ▶

| | | | |
|---|---|---|---|
| Lymphatic vessel endothelial hyaluronic acid receptor 1 (LYVE1) | Q9Y5Y7 | Plexin-B2 (PLXNB2) | O15031 |
| Lysosomal Pro-X carboxypeptidase (PRCP) | P42785 | Procollagen C-endopeptidase enhancer 1 (PCOLCE) | Q15113 |
| Mannose-binding protein C (MBL2) | P11226 | Prolyl endopeptidase FAP (FAP) | Q12884 |
| Mast/stem cell growth factor receptor Kit (KIT) | P10721 | Receptor-type tyrosine-protein phosphatase S (PTPRS) | Q13332 |
| Membrane cofactor protein (CD46) | P15529 | Regenerating islet-derived protein 3-alpha (REG3A) | Q06141 |
| Membrane primary amine oxidase (AOC3) | Q16853 | Serum amyloid A-4 protein (SAA4) | P35542 |
| Metalloproteinase inhibitor 1 (TIMP1) | P01033 | SPARC-like protein 1 (SPARCL1) | Q14515 |
| Microfibrillar-associated protein 5 (MFAP5) | Q13361 | Superoxide dismutase [Cu-Zn] (SOD1) | P00441 |
| Multiple epidermal growth factor-like domains protein 9 (MEGF9) | Q9H1U4 | T-cell immunoglobulin and mucin domain-containing protein 4 (TIMD4) | Q96H15 |
| Neural cell adhesion molecule 1 (NCAM1) | P13591 | Tenascin (TNC) | P24821 |
| Neural cell adhesion molecule L1-like protein (CHL1) | O00533 | Tenascin-X (TNXB) | P22105 |
| Neurogenic locus notch homolog protein 1 (NOTCH1) | P46531 | Thrombospondin-4 (THBS4) | P35443 |
| Neuropilin-1 (NRP1) | O14786 | Thyroxine-binding globulin (SERPINA7) | P05543 |
| Neutrophil defensin 1 (DEFA1) | P59665 | Transcobalamin-2 (TCN2) | P20062 |
| Neutrophil gelatinase-associated lipocalin (LCN2) | P80188 | Transforming growth factor beta receptor type 3 (TGFBR3) | Q03167 |
| Nidogen-1 (NID1) | P14543 | Transforming growth factor-beta-induced protein ig-h3 (TGFBI) | Q15582 |
| Oncostatin-M-specific receptor subunit beta (OSMR) | Q99650 | Trypsin-2 (PRSS2) | P07478 |
| Peptidyl-glycine alpha-amidating monooxygenase (PAM) | P19021 | Tyrosine-protein kinase receptor Tie-1 (TIE1) | P35590 |
| Phospholipid transfer protein (PLTP) | P55058 | Uromodulin (UMOD) | P07911 |
| Plasma serine protease inhibitor (SERPINA5) | P05154 | Vascular cell adhesion protein 1 (VCAM1) | P19320 |
| Platelet glycoprotein Ib alpha chain (GP1BA) | P07359 | Vasorin (VASN) | Q6EMK4 |
| Platelet-activating factor acetylhydrolase (PLA2G7) | Q13093 | Vitamin K-dependent protein C (PROC) | P04070 |

For more details visit www.olink.com/cardiometabolic

### www.olink.com

For research use only. Not for use in diagnostic procedures.
This product includes a license for non-commercial use. Commercial users may require additional licenses. Please contact Olink Proteomics AB for details.
There are no warranties, expressed or implied, which extend beyond this description. Olink Proteomics AB is not liable for property damage, personal injury, or economic loss caused by this product.
Olink® is a registered trademark of Olink Proteomics AB.

© 2017–2022 Olink Proteomics AB. All third party trademarks are the property of their respective owners.

Olink Proteomics, Dag Hammarskjölds väg 52B , SE-752 37 Uppsala, Sweden
1065, v2.0, 2022-06-14

Olink®

Protein assay list

### Olink® Target 96 Cell Regulation

Product number: 95370

| | | | |
|---|---|---|---|
| Amphoterin-induced protein 2 (AMIGO2) | Q86SJ2 | Discoidin, CUB and LCCL domain-containing protein 2 (DCBLD2) | Q96PD2 |
| Amyloid beta A4 precursor protein-binding family B member 1-interacting protein (APBB1IP) | Q7Z5R6 | DnaJ homolog subfamily B member 1 (DNAJB1) | P25685 |
| | | Dual specificity mitogen-activated protein kinase kinase 6 (MAP2K6) | P52564 |
| Anterior gradient protein 3 (AGR3) | Q8TD06 | Dynactin subunit 2 (DCTN2) | Q13561 |
| Arylsulfatase B (ARSB) | P15848 | E3 ubiquitin-protein ligase CBL (CBL) | P22681 |
| ATP-dependent 6-phosphofructokinase, muscle type (PFKM) | P08237 | Ectonucleoside triphosphate diphosphohydrolase 6 (ENTPD6) | O75354 |
| Bcl-2-like protein 11 isoform BimL (BCL2L11) | O43521 | Fibroblast growth factor 21 (FGF21) | Q9NSA1 |
| Beta-1,3-galactosyl-O-glycosyl-glycoprotein beta-1,6-N-acetylglucosaminyltransferase (GCNT1) | Q02742 | Friend leukemia integration 1 transcription factor (FLI1) | Q01543 |
| Biglycan (BGN) | P21810 | Galectin-7 (LGALS7) | P47929 |
| Bile salt sulfotransferase (SULT2A1) | Q06520 | Gamma-secretase-activating protein (GSAP) | A4D1B5 |
| Breakpoint cluster region protein (BCR) | P11274 | Gastrokine (GKN1) | Q9NS71 |
| Brother of CDO (BOC) | Q9BWV1 | GDNF family receptor alpha-2 (GFRA2) | O00451 |
| Calcium/calmodulin-dependent protein kinase kinase 1 (CAMKK1) | Q8N5S9 | Glucagon (GCG) | P01275 |
| Calsyntenin-3 (CLSTN3) | Q9BQT9 | Growth hormone variant (GH2) | P01242 |
| Cell growth-regulating nucleolar protein (LYAR) | Q9NX58 | Heparan sulfate glucosamine 3-O-sulfotransferase 3B1 (HS3ST3B1) | Q9Y662 |
| Cellular tumor antigen p53 (TP53) | P04637 | Heparan-sulfate 6-O-sulfotransferase 1 (HS6ST1) | O60243 |
| Cerebral dopamine neurotrophic factor (CDNF) | Q49AH0 | Immunoglobulin superfamily member 3 (IGSF3) | O75054 |
| Collagen alpha-1(IV) chain (COL4A1) | P02462 | Interleukin-17 receptor B (IL17RB) | Q9NRM6 |
| Cone-rod homeobox protein (CRX) | O43186 | Kallikrein-12 (KLK12) | Q9UKR0 |
| Cryptic protein (CFC1) | P0CG37 | Kazal-type serine protease inhibitor domain-containing protein 1 (KAZALD1) | Q96I82 |
| C-type mannose receptor 2 (MRC2) | Q9UBG0 | | |
| Cysteine protease ATG4A (ATG4A) | Q8WYN0 | Leucine-rich repeat neuronal protein 1 (LRRN1) | Q6UXK5 |
| Cysteine-rich secretory protein 2 (CRISP2) | P16562 | Ly6/PLAUR domain-containing protein 1 (LYPD1) | Q8N2G4 |
| Dickkopf-like protein 1 (DKKL1) | Q9UK85 | Lymphoid-restricted membrane protein (LRMP) | Q12912 |

Table continues on reverse ▶

| | | | |
|---|---|---|---|
| Methionine aminopeptidase 1D, mitochondrial (METAP1D) | Q6UB28 | Semaphorin-4C (SEMA4C) | Q9C0C4 |
| Myelin-oligodendrocyte glycoprotein (MOG) | Q16653 | Serine/threonine-protein kinase PAK 4 (PAK4) | O96013 |
| N(G),N(G)-dimethylarginine dimethylaminohydrolase 1 (DDAH1) | O94760 | Sialic acid-binding Ig-like lectin 10 (SIGLEC10) | Q96LC7 |
| NF-kappa-B inhibitor epsilon (NFKBIE) | O00221 | Sialic acid-binding Ig-like lectin 6 (SIGLEC6) | O43699 |
| Nicalin (NCLN) | Q969V3 | SLAM family member 8 (SLAMF8) | Q9P0V8 |
| Ninjurin-1 (NINJ1) | Q92982 | SLIT and NTRK-like protein 2 (SLITRK2) | Q9H156 |
| Nuclear factor of activated T-cells, cytoplasmic 1 (NFATC1) | O95644 | SLIT and NTRK-like protein 6 (SLITRK6) | Q9H5Y7 |
| Oligodendrocyte-myelin glycoprotein (OMG) | P23515 | Src kinase-associated phosphoprotein 1 (SKAP1) | Q86WV1 |
| Opticin (OPTC) | Q9UBM4 | Syntaxin-6 (STX6) | O43752 |
| Peroxiredoxin-6 (PRDX6) | P30041 | Syntaxin-16 (STX16) | O14662 |
| Podocalyxin-like protein 2 (PODXL2) | Q9NZ53 | T-cell leukemia/lymphoma protein 1B (TCL1B) | O95988 |
| Polypeptide N-acetylgalactosaminyltransferase 2 (GALNT2) | Q10471 | Transcription factor AP-1 (JUN) | P05412 |
| Probable carboxypeptidase X1 (CPXM1) | Q96SM3 | Transforming acidic coiled-coil-containing protein 3 (TACC3) | Q9Y6A5 |
| Prokineticin-1 (PROK1) | P58294 | Tudor and KH domain-containing protein (TDRKH) | Q9Y2W6 |
| Prolactin regulatory element-binding protein (PREB) | Q9HCU5 | Tumor necrosis factor receptor superfamily member 10A (TNFRSF10A) | O00220 |
| Protein FAM19A5 (FAM19A5) | Q7Z5A7 | Tumor-associated calcium signal transducer 2 (TACSTD2) | P09758 |
| Protein Wnt-9a (WNT9A) | O14904 | Vascular endothelial growth factor D (VEGFD) | O43915 |
| Protocadherin-17 (PCDH17) | O14917 | Vesicle-associated membrane protein 5 (VAMP5) | O95183 |
| Ras GTPase-activating-like protein IQGAP2 (IQGAP2) | Q13576 | VPS10 domain-containing receptor SorCS2 (SORCS2) | Q96PQ0 |
| Regulator of G-protein signaling 8 (RGS8) | P57771 | Wiskott-Aldrich syndrome protein family member 1 (WASF1) | Q92558 |
| Rho GTPase-activating protein 1 (ARHGAP1) | Q07960 | Wiskott-Aldrich syndrome protein family member 3 (WASF3) | Q9UPY6 |
| Rho guanine nucleotide exchange factor 12 (ARHGEF12) | Q9NZN5 | Zinc finger and BTB domain-containing protein 16 (ZBTB16) | Q05516 |
| Seizure 6-like protein 2 (SEZ6L2) | Q6UXD5 | Zinc finger and BTB domain-containing protein 17 (ZBTB17) | Q13105 |

For more details visit www.olink.com/cell-regulation

www.olink.com

For research use only. Not for use in diagnostic procedures.
This product includes a license for non-commercial use. Commercial users may require additional licenses. Please contact Olink Proteomics AB for details.
There are no warranties, expressed or implied, which extend beyond this description. Olink Proteomics AB is not liable for property damage, personal injury, or economic loss caused by this product.
Olink® is a registered trademark of Olink Proteomics AB.

© 2017–2022 Olink Proteomics AB. All third party trademarks are the property of their respective owners.
Olink Proteomics, Dag Hammarskjölds väg 52B , SE-752 37 Uppsala, Sweden
1066, v2.0, 2022-06-14

Olink®

Protein assay list

### Olink® Target 96 Cardiovascular II

Product number: 95500

| | | | |
|---|---|---|---|
| 2,4-dienoyl-CoA reductase, mitochondrial (DECR1) | Q16698 | Gastric intrinsic factor (GIF) | P27352 |
| A disintegrin and metalloproteinase with thrombospondin motifs 13 (ADAM-TS13) | Q76LX8 | Gastrotropin (GT) | P51161 |
| ADM (ADM) | P35318 | Growth hormone (GH) | P01241 |
| Agouti-related protein (AGRP) | O00253 | Growth/differentiation factor 2 (GDF-2) | Q9UK05 |
| Alpha-L-iduronidase (IDUA) | P35475 | Heat shock 27 kDa protein (HSP 27) | P04792 |
| Angiopoietin-1 (ANG-1) | Q15389 | Heme oxygenase 1 (HO-1) | P09601 |
| Angiopoietin-1 receptor (TIE2) | Q02763 | Hydroxyacid oxidase 1 (HAOX1) | Q9UJM8 |
| Angiotensin-converting enzyme 2 (ACE2) | Q9BYF1 | Interleukin-1 receptor antagonist protein (IL-1ra) | P18510 |
| Bone morphogenetic protein 6 (BMP-6) | P22004 | Interleukin-1 receptor-like 2 (IL1RL2) | Q9HB29 |
| Brother of CDO (Protein BOC) | Q9BWV1 | Interleukin-4 receptor subunit alpha (IL-4RA) | P24394 |
| Carbonic anhydrase 5A, mitochondrial (CA5A) | P35218 | Interleukin-6 (IL6) | P05231 |
| Carcinoembryonic antigenrelated cell adhesion molecule 8 (CEACAM8) | P31997 | Interleukin-17D (IL-17D) | Q8TAD2 |
| Cathepsin L1 (CTSL1) | P07711 | Interleukin-18 (IL-18) | Q14116 |
| C-C motif chemokine 3 (CCL3) | P10147 | Interleukin-27 (IL-27) | Q8NEV9, Q14213 |
| C-C motif chemokine 17 (CCL17) | Q92583 | Kidney Injury Molecule (KIM1) | Q96D42 |
| CD40 ligand (CD40-L) | P29965 | Lactoylglutathione lyase (GLO1) | Q04760 |
| Chymotrypsin C (CTRC) | Q99895 | Lectin-like oxidized LDL receptor 1 (LOX-1) | P78380 |
| C-X-C motif chemokine 1 (CXCL1) | P09341 | Leptin (LEP) | P41159 |
| Decorin (DCN) | P07585 | Lipoprotein lipase (LPL) | P06858 |
| Dickkopf-related protein 1 (Dkk-1) | O94907 | Low affinity immunoglobulin gamma Fc region receptor II-b (IgG Fc receptor II-b) | P31994 |
| Fatty acid-binding protein, intestinal (FABP2) | P12104 | Lymphotactin (XCL1) | P47992 |
| Fibroblast growth factor 21 (FGF-21) | Q9NSA1 | Macrophage receptor MARCO (MARCO) | Q9UEW3 |
| Fibroblast growth factor 23 (FGF-23) | Q9GZV9 | Matrix metalloproteinase-7 (MMP-7) | P09237 |
| Follistatin (FS) | P19883 | Matrix metalloproteinase-12 (MMP-12) | P39900 |
| Galectin-9 (Gal-9) | O00182 | Melusin (ITGB1BP2) | Q9UKP3 |

Table continues on reverse ▶

| | | | |
|---|---|---|---|
| Natriuretic peptides B (BNP) | P16860 | Serine protease 27 (PRSS27) | Q9BQR3 |
| NF-kappa-B essential modulator (NEMO) | Q9Y6K9 | Serine/threonine-protein kinase 4 (STK4) | Q13043 |
| Osteoclast-associated immunoglobulin-like receptor (hOSCAR) | Q8IYS5 | Serpin A12 (SERPINA12) | Q8IW75 |
| Pappalysin-1 (PAPPA) | Q13219 | SLAM family member 5 (CD84) | Q9UIB8 |
| Pentraxin-related protein PTX3 (PTX3) | P26022 | SLAM family member 7 (SLAMF7) | Q9NQ25 |
| Placenta growth factor (PGF) | P49763 | Sortilin (SORT1) | Q99523 |
| Platelet-derived growth factor subunit B (PDGF subunit B) | P01127 | Spondin-2 (SPON2) | Q9BUD6 |
| Poly [ADP-ribose] polymerase 1 (PARP-1) | P09874 | Stem cell factor (SCF) | P21583 |
| Polymeric immunoglobulin receptor (PIgR) | P01833 | Superoxide dismutase [Mn], mitochondrial (SOD2) | P04179 |
| Programmed cell death 1 ligand 2 (PD-L2) | Q9BQ51 | T-cell surface glycoprotein CD4 (CD4) | P01730 |
| Proheparin-binding EGF-like growth factor (HB-EGF) | Q99075 | Thrombomodulin TM | P07204 |
| Pro-interleukin-16 (IL16) | Q14005 | Thrombopoietin (THPO) | P40225 |
| Prolargin (PRELP) | P51888 | Thrombospondin-2 (THBS2) | P35442 |
| Prostasin (PRSS8 ) | Q16651 | Tissue factor (TF) | P13726 |
| Protein AMBP (AMBP) | P02760 | TNF-related apoptosis-inducing ligand receptor 2 (TRAIL-R2) | O14763 |
| Proteinase-activated receptor 1 (PAR-1) | P25116 | Tumor necrosis factor receptor superfamily member 10A (TNFRSF10A) | O00220 |
| Protein-glutamine gamma-glutamyltransferase 2 (TGM2) | P21980 | Tumor necrosis factor receptor superfamily member 11A (TNFRSF11A) | Q9Y6Q6 |
| Proto-oncogene tyrosine-protein kinase Src (SRC) | P12931 | Tumor necrosis factor receptor superfamily member 13B (TNFRSF13B) | O14836 |
| P-selectin glycoprotein ligand 1 (PSGL-1) | Q14242 | Tyrosine-protein kinase Mer (MERTK) | Q12866 |
| Receptor for advanced glycosylation end products (RAGE) | Q15109 | Vascular endothelial growth factor D (VEGFD) | O43915 |
| Renin (REN) | P00797 | V-set and immunoglobulin domain-containing protein 2 (VSIG2) | Q96IQ7 |

For more details visit www.olink.com/cvd2

### www.olink.com

For research use only. Not for use in diagnostic procedures.
This product includes a license for non-commercial use. Commercial users may require additional licenses. Please contact Olink Proteomics AB for details.
There are no warranties, expressed or implied, which extend beyond this description. Olink Proteomics AB is not liable for property damage, personal injury, or economic loss caused by this product.
Olink® is a registered trademark of Olink Proteomics AB.

© 2017–2022 Olink Proteomics AB. All third party trademarks are the property of their respective owners.
Olink Proteomics, Dag Hammarskjölds väg 52B , SE-752 37 Uppsala, Sweden
1024, v2.0, 2022-06-14

Olink®

Protein assay list

### Olink® Target 96 Cardiovascular III

Product number: 95611

| | | | |
|---|---|---|---|
| Aminopeptidase N (AP-N) | P15144 | Galectin-3 (Gal-3) | P17931 |
| Azurocidin (AZU1) | P20160 | Galectin-4 (Gal-4) | P56470 |
| Bleomycin hydrolase (BLM hydrolase) | Q13867 | Granulins (GRN) | P28799 |
| Cadherin-5 (CDH5) | P33151 | Growth/differentiation factor 15 (GDF-15) | Q99988 |
| Carboxypeptidase A1 (CPA1) | P15085 | Insulin-like growth factor-binding protein 1 (IGFBP-1) | P08833 |
| Carboxypeptidase B (CPB1) | P15086 | Insulin-like growth factor-binding protein 2 (IGFBP-2) | P18065 |
| Caspase-3 (CASP-3) | P42574 | Insulin-like growth factor-binding protein 7 (IGFBP-7) | Q16270 |
| Cathepsin D (CTSD) | P07339 | Integrin beta-2 (ITGB2) | P05107 |
| Cathepsin Z (CTSZ) | Q9UBR2 | Intercellular adhesion molecule 2 (ICAM-2) | P13598 |
| C-C motif chemokine 15 (CCL15) | Q16663 | Interleukin-1 receptor type 1 (IL-1RT1) | P14778 |
| C-C motif chemokine 16 (CCL16) | O15467 | Interleukin-1 receptor type 2 (IL-1RT2) | P27930 |
| C-C motif chemokine 24 (CCL24) | O00175 | Interleukin-2 receptor subunit alpha (IL2-RA) | P01589 |
| CD166 antigen (ALCAM) | Q13740 | Interleukin-6 receptor subunit alpha (IL-6RA) | P08887 |
| Chitinase-3-like protein 1 (CHI3L1) | P36222 | Interleukin-17 receptor A (IL-17RA) | Q96F46 |
| Chitotriosidase-1 (CHIT1) | Q13231 | Interleukin-18-binding protein (IL-18BP) | O95998 |
| Collagen alpha-1(I) chain (COL1A1) | P02452 | Junctional adhesion molecule A (JAM-A) | Q9Y624 |
| Complement component C1q receptor (CD93) | Q9NPY3 | Kallikrein-6 (KLK6) | Q92876 |
| Contactin-1 (CNTN1) | Q12860 | Low-density lipoprotein receptor (LDL receptor) | P01130 |
| C-X-C motif chemokine 16 (CXCL16) | Q9H2A7 | Lymphotoxin-beta receptor (LTBR) | P36941 |
| Cystatin-B (CSTB) | P04080 | Matrix extracellular phosphoglycoprotein (MEPE) | Q9NQ76 |
| Elafin (PI3) | P19957 | Matrix metalloproteinase-2 (MMP-2) | P08253 |
| Ephrin type-B receptor 4 (EPHB4) | P54760 | Matrix metalloproteinase-3 (MMP-3) | P08254 |
| Epidermal growth factor receptor (EGFR) | P00533 | Matrix metalloproteinase-9 (MMP-9) | P14780 |
| Epithelial cell adhesion molecule (Ep-CAM) | P16422 | Metalloproteinase inhibitor 4 (TIMP4) | Q99727 |
| E-selectin (SELE) | P16581 | Monocyte chemotactic protein 1 (MCP-1) | P13500 |
| Fatty acid-binding protein, adipocyte (FABP4) | P15090 | Myeloblastin (PRTN3) | P24158 |

Table continues on reverse ▶

| | | | |
|---|---|---|---|
| Myeloperoxidase (MPO) | P05164 | Secretoglobin family 3A member 2 (SCGB3A2) | Q96PL1 |
| Myoglobin (MB) | P02144 | Spondin-1 (SPON1) | Q9HCB6 |
| Neurogenic locus notch homolog protein 3 (Notch 3) | Q9UM47 | ST2 protein (ST2) | Q01638 |
| N-terminal prohormone brain natriuretic peptide (NT-proBNP) | NA | Tartrate-resistant acid phosphatase type 5 (TR-AP) | P13686 |
| Osteopontin (OPN) | P10451 | Tissue factor pathway inhibitor (TFPI) | P10646 |
| Osteoprotegerin (OPG) | O00300 | Tissue-type plasminogen activator (t-PA) | P00750 |
| Paraoxonase (PON3) | Q15166 | Transferrin receptor protein 1 (TR) | P02786 |
| Peptidoglycan recognition protein 1 (PGLYRP1) | O75594 | Trefoil factor 3 (TFF3) | Q07654 |
| Perlecan (PLC) | P98160 | Trem-like transcript 2 protein (TLT-2) | Q5T2D2 |
| Plasminogen activator inhibitor 1 (PAI) | P05121 | Tumor necrosis factor ligand superfamily member 13B (TNFSF13B) | Q9Y275 |
| Platelet endothelial cell adhesion molecule (PECAM-1) | P16284 | Tumor necrosis factor receptor 1 (TNF-R1) | P19438 |
| Platelet-derived growth factor subunit A (PDGF subunit A) | P04085 | Tumor necrosis factor receptor 2 (TNF-R2) | P20333 |
| Platelet glycoprotein VI (GP6) | Q9HCN6 | Tumor necrosis factor receptor superfamily member 6 (FAS) | P25445 |
| Proprotein convertase subtilisin/kexin type 9 (PCSK9) | Q8NBP7 | Tumor necrosis factor receptor superfamily member 10C (TNFRSF10C) | O14798 |
| Protein delta homolog 1 (DLK-1) | P80370 | Tumor necrosis factor receptor superfamily member 14 (TNFRSF14) | Q92956 |
| P-selectin (SELP) | P16109 | Tyrosine-protein kinase receptor UFO (AXL) | P30530 |
| Pulmonary surfactant-associated protein D (PSP-D) | P35247 | Tyrosine-protein phosphatase non-receptor type substrate 1 (SHPS-1) | P78324 |
| Resistin (RETN) | Q9HD89 | Urokinase plasminogen activator surface receptor (U-PAR) | Q03405 |
| Retinoic acid receptor responder protein 2 (RARRES2) | Q99969 | Urokinase-type plasminogen activator (uPA) | P00749 |
| Scavenger receptor cysteine-rich type 1 protein M130 (CD163) | Q86VB7 | von Willebrand factor (vWF) | P04275 |

For more details visit www.olink.com/cvd3

### www.olink.com

For research use only. Not for use in diagnostic procedures.
This product includes a license for non-commercial use. Commercial users may require additional licenses. Please contact Olink Proteomics AB for details.
There are no warranties, expressed or implied, which extend beyond this description. Olink Proteomics AB is not liable for property damage, personal injury, or economic loss caused by this product.
Olink® is a registered trademark of Olink Proteomics AB.

© 2017–2022 Olink Proteomics AB. All third party trademarks are the property of their respective owners.

Olink Proteomics, Dag Hammarskjölds väg 52B , SE-752 37 Uppsala, Sweden
1023, v2.0, 2022-06-03

Olink®

Protein assay list

### Olink® Target 96 Development

Product number: 95352

| | | | |
|---|---|---|---|
| ADP-ribosyl cyclase/cyclic ADP-ribose hydrolase 2 (BST1) | Q10588 | Contactin-4 (CNTN4) | Q8IWV2 |
| ADP-sugar pyrophosphatase (NUDT5) | Q9UKK9 | Corticotropin-releasing factor-binding protein (CRHBP) | P24387 |
| Aggrecan core protein (ACAN) | P16112 | C-type lectin domain family 11 member A (CLEC11A) | Q9Y240 |
| Amyloid beta A4 protein (APP) | P05067 | C-type lectin domain family 14 member A (CLEC14A) | Q86T13 |
| Angiopoietin-related protein 4 (ANGPTL4) | Q9BY76 | Cystatin-M (CST6) | Q15828 |
| Arylsulfatase A (ARSA) | P15289 | Cysteine-rich motor neuron 1 protein (CRIM1) | Q9NZV1 |
| Basal cell adhesion molecule (BCAM) | P50895 | Cysteine-rich with EGF-like domain protein 2 (CRELD2) | Q6UXH1 |
| Beta-1,4-galactosyltransferase 1 (B4GALT1) | P15291 | Cytosolic phospholipase A2 (PLA2G4A) | P47712 |
| Beta-1,4-glucuronyltransferase 1 (B4GAT1) | O43505 | Desmocollin-2 (DSC2) | Q02487 |
| Beta-glucuronidase (GUSB) | P08236 | Dickkopf-related protein 3 (DKK3) | Q9UBP4 |
| Beta-microseminoprotein (MSMB) | P08118 | Dystroglycan (DAG1) | Q14118 |
| Carbonic anhydrase 2 (CA2) | P00918 | Early activation antigen CD69 (CD69) | Q07108 |
| Carbonic anhydrase 6 (CA6) | P23280 | Ectonucleotide pyrophosphatase/phosphodiesterase family member 2 (ENPP2) | Q13822 |
| Cathepsin F (CTSF) | Q9UBX1 | Endothelial cell-selective adhesion molecule (ESAM) | Q96AP7 |
| Cation-independent mannose-6-phosphate receptor (IGF2R) | P11717 | Fc receptor-like protein 5 (FCRL5) | Q96RD9 |
| C-C motif chemokine 21 (CCL21) | O00585 | Flavin reductase NADPH (BLVRB) | P30043 |
| CCN family member 5 (CCN5) | O76076 | Follistatin-related protein 3 (FSTL3) | O95633 |
| CD97 antigen (CD97) | P48960 | Galactoside 3(4)-L-fucosyltransferase, Alpha-(1,3)-fucosyltransferase 3/5 (FUT3/5) | P21217, Q11128 |
| CD99 antigen-like protein 2 (CD99L2) | Q8TCZ2 | Glycoprotein hormones alpha chain (CGA) | P01215 |
| CD109 antigen (CD109) | Q6YHK3 | Glycoprotein Xg (XG) | P55808 |
| CD177 antigen (CD177) | Q8N6Q3 | Hepatitis A virus cellular receptor 2 (HAVCR2) | Q8TDQ0 |
| CD209 antigen (CD209) | Q9NNX6 | HLA class II histocompatibility antigen gamma chain (CD74) | P04233 |
| Cell adhesion molecule-related/down-regulated by oncogenes (CDON) | Q4KMG0 | Inactive serine protease PAMR1 (PAMR1) | Q6UXH9 |
| CMRF35-like molecule 9 (CD300LG) | Q6UXG3 | Inhibin beta C chain (INHBC) | P55103 |
| Cochlin (COCH) | O43405 | Integrin alpha-5 (ITGA5) | P08648 |
| Collectin-12 (COLEC12) | Q5KU26 | Integrin beta-1 (ITGB1) | P05556 |

Table continues on reverse ▶

| | |
|---|---|
| Interleukin-13 receptor subunit alpha-1 (IL13RA1) | P78552 |
| Kunitz-type protease inhibitor 1 (SPINT1) | O43278 |
| Kunitz-type protease inhibitor 2 (SPINT2) | O43291 |
| Lactadherin (MFGE8) | Q08431 |
| Laminin subunit alpha-4 (LAMA4) | Q16363 |
| LDLR chaperone MESD (MESDC2) | Q14696 |
| Legumain (LGMN) | Q99538 |
| Leukocyte-associated immunoglobulin-like receptor 1 (LAIR1) | Q6GTX8 |
| Low affinity immunoglobulin epsilon Fc receptor (FCER2) | P06734 |
| Lymphocyte function-associated antigen 3 (CD58) | P19256 |
| Macrophage migration inhibitory factor (MIF) | P14174 |
| Matrilin-2 (MATN2) | O00339 |
| Myocilin (MYOC) | Q99972 |
| Nidogen-2 (NID2) | Q14112 |
| Osteomodulin (OMD) | Q99983 |
| Paired immunoglobulin-like type 2 receptor alpha (PILRA) | Q9UKJ1 |
| Peptidyl-prolyl cis-trans isomerase B (PPIB) | P23284 |
| Phosphatidylethanolamine-binding protein 1 (PEBP1) | P30086 |
| Platelet endothelial aggregation receptor 1 (PEAR1) | Q5VY43 |
| Platelet-derived growth factor receptor beta (PDGFRB) | P09619 |
| Protein deglycase DJ-1 (PARK7) | Q99497 |
| Protein disulfide-isomerase (P4HB) | P07237 |
| Protein NOV homolog (NOV) | P48745 |
| Receptor-type tyrosine-protein phosphatase F (PTPRF) | P10586 |
| Roundabout homolog 1 (ROBO1) | Q9Y6N7 |
| Scavenger receptor class F member 1 (SCARF1) | Q14162 |
| Semaphorin-7A (SEMA7A) | O75326 |
| Serine protease HTRA2, mitochondrial (HTRA2) | O43464 |
| Serine protease inhibitor Kazal-type 1 (SPINK1) | P00995 |
| Serine protease inhibitor Kazal-type 5 (SPINK5) | Q9NQ38 |
| Signal-regulatory protein beta-1 (SIRPB1) | O00241 |
| Stress-induced-phosphoprotein 1 (STIP1) | P31948 |
| Synaptosomal-associated protein 29 (SNAP29) | O95721 |
| Thymosin beta-10 (TMSB10) | P63313 |
| Tissue alpha-L-fucosidase (FUCA1) | P04066 |
| Tripeptidyl-peptidase 1 (TPP1) | O14773 |
| Tumor necrosis factor receptor superfamily member 19L (RELT) | Q969Z4 |
| Tyrosine-protein phosphatase non-receptor type 6 (PTPN6) | P29350 |
| WAP, Kazal, immunoglobulin, Kunitz and NTR domain-containing protein 2 (WFIKKN2) | Q8TEU8 |
| V-set and immunoglobulin domain-containing protein 4 (VSIG4) | Q9Y279 |

For more details visit www.olink.com/development

www.olink.com

For research use only. Not for use in diagnostic procedures.
This product includes a license for non-commercial use. Commercial users may require additional licenses. Please contact Olink Proteomics AB for details.
There are no warranties, expressed or implied, which extend beyond this description. Olink Proteomics AB is not liable for property damage, personal injury, or economic loss caused by this product.
Olink® is a registered trademark of Olink Proteomics AB.

© 2017–2022 Olink Proteomics AB. All third party trademarks are the property of their respective owners.

Olink Proteomics, Dag Hammarskjölds väg 52B , SE-752 37 Uppsala, Sweden
1064, v2.0, 2022-06-14

Protein assay list

### Olink® Target 96 Immune Response

Product number: 95320

| | | | |
|---|---|---|---|
| Allergin-1 (MILR1) | Q7Z6M3 | Eotaxin (CCL11) | P51671 |
| Amphiregulin (AR) (AREG) | P15514 | Eukaryotic translation initiation factor 4 gamma 1 (EIF4G1) | Q04637 |
| Aryl hydrocarbon receptor nuclear translocator (ARNT) | P27540 | Eukaryotic translation initiation factor 5A-1 (EIF5A) | P63241 |
| Baculoviral IAP repeat-containing protein 2 (BIRC2) | Q13490 | Fc receptor-like protein 3 (FCRL3) | Q96P31 |
| Beta-galactosidase (GLB1) | P16278 | Fc receptor-like protein 6 (FCRL6) | Q6DN72 |
| Butyrophilin subfamily 3 member A2 (BTN3A2) | P78410 | Fibroblast growth factor 2 (FGF2) | P09038 |
| CD83 antigen (CD83) | Q01151 | FXYD domain-containing ion transport regulator 5 (FXYD5) | Q96DB9 |
| Contactin-associated protein-like 2 (CNTNAP2) | Q9UHC6 | Hematopoietic lineage cell-specific protein (HCLS1) | P14317 |
| Corneodesmosin (CDSN) | Q15517 | Histamine N-methyltransferase (HNMT) | P50135 |
| Corticosteroid 11-beta-dehydrogenase isozyme 1 (HSD11B1) | P28845 | Importin subunit alpha-5 (KPNA1) | P52294 |
| Coxsackievirus and adenovirus receptor (CXADR) | P78310 | Inactive dipeptidyl peptidase 10 (DPP10) | Q8N608 |
| C-type lectin domain family 4 member A (CLEC4A) | Q9UMR7 | Integral membrane protein 2A (ITM2A) | O43736 |
| C-type lectin domain family 4 member C (CLEC4C) | Q8WTT0 | Integrin alpha-6 (ITGA6) | P23229 |
| C-type lectin domain family 4 member D (CLEC4D) | Q8WXI8 | Integrin alpha-11 (ITGA11) | Q9UKX5 |
| C-type lectin domain family 4 member G (CLEC4G) | Q6UXB4 | Integrin beta-6 (ITGB6) | P18564 |
| C-type lectin domain family 6 member A (CLEC6A) | Q6EIG7 | Interferon lambda receptor 1 (IFNLR1) | Q8IU57 |
| C-type lectin domain family 7 member A (CLEC7A) | Q9BXN2 | Interferon regulatory factor 9 (IRF9) | Q00978 |
| Cytoskeleton-associated protein 4 (CKAP4) | Q07065 | Interleukin-1 receptor-associated kinase 1 (IRAK1) | P51617 |
| Diacylglycerol kinase zeta (DGKZ) | Q13574 | Interleukin-1 receptor-associated kinase 4 (IRAK4) | Q9NWZ3 |
| Discoidin, CUB and LCCL domain-containing protein 2 (DCBLD2) | Q96PD2 | Interleukin-5 (IL5) | P05113 |
| DNA fragmentation factor subunit alpha (DFFA) | O00273 | Interleukin-6 (IL6) | P05231 |
| Dual adapter for phosphotyrosine and 3-phosphotyrosine and 3-phosphoinositide (DAPP1) | Q9UN19 | Interleukin-10 (IL10) | P22301 |
| Dynactin subunit 1 (DCTN1) | Q14203 | Interleukin-12 receptor subunit beta-1 (IL12RB1) | P42701 |
| E3 ubiquitin-protein ligase TRIM21 (TRIM21) | P19474 | Islet cell autoantigen 1 (ICA1) | Q05084 |
| Egl nine homolog 1 (EGLN1) | Q9GZT9 | Keratin, type I cytoskeletal 19 (KRT19) | P08727 |

Table continues on reverse ▶

| | | | |
|---|---|---|---|
| Leukocyte immunoglobulin-like receptor subfamily B member 4 (LILRB4) | Q8NHJ6 | Protein HEXIM1 (HEXIM1) | O94992 |
| Lymphocyte activation gene 3 protein (LAG3) | P18627 | Protein kinase C theta type (PRKCQ) | Q04759 |
| Lymphocyte antigen 75 (LY75) | O60449 | Protein sprouty homolog 2 (SPRY2) | O43597 |
| Lysosome-associated membrane glycoprotein 3 (LAMP3) | Q9UQV4 | Protein-arginine deiminase type-2 (PADI2) | Q9Y2J8 |
| Mannan-binding lectin serine protease 1 (MASP1) | P48740 | SH2 domain-containing protein 1A (SH2D1A) | O60880 |
| Merlin (NF2) | P35240 | SH2B adapter protein 3 (SH2B3) | Q9UQQ2 |
| Methylated-DNA--protein-cysteine methyltransferase (MGMT) | P16455 | Signaling threshold-regulating transmembrane adapter 1 (SIT1) | Q9Y3P8 |
| Natural cytotoxicity triggering receptor 1 (NCR1) | O76036 | SRSF protein kinase 2 (SRPK2) | P78362 |
| Natural killer cells antigen CD94 (KLRD1) | Q13241 | Stanniocalcin-1 (STC1) | P52823 |
| Neurabin-2 (PPP1R9B) | Q96SB3 | Stromal cell-derived factor 1 (CXCL12) | P48061 |
| Neurotrophin-4 (NTF4) | P34130 | T-cell-specific surface glycoprotein CD28 (CD28) | P10747 |
| Nuclear factor of activated T-cells, cytoplasmic 3 (NFATC3) | Q12968 | Thioredoxin-dependent peroxide reductase, mitochondrial (PRDX3) | P30048 |
| Parathyroid hormone/parathyroid hormone-related peptide receptor (PTH1R) | Q03431 | TNF receptor-associated factor 2 (TRAF2) | Q12933 |
| PC4 and SFRS1-interacting protein (PSIP1) | O75475 | TRAF family member-associated NF-kappa-B activator (TANK) | Q92844 |
| Peroxiredoxin-1 (PRDX1) | Q06830 | Transcription factor AP-1 (JUN) | P05412 |
| Peroxiredoxin-5, mitochondrial (PRDX5) | P30044 | Transcription regulator protein BACH1 (BACH1) | O14867 |
| Phosphoinositide 3-kinase adapter protein 1 (PIK3AP1) | Q6ZUJ8 | Triggering receptor expressed on myeloid cells 1 (TREM1) | Q9NP99 |
| Plexin-A4 (PLXNA4) | Q9HCM2 | Tripartite motif-containing protein 5 (TRIM5) | Q9C035 |
| Polypeptide N-acetylgalactosaminyltransferase 3 (GALNT3) | Q14435 | Tryptase alpha/beta-1 (TPSAB1) | Q15661 |
| Probable ATP-dependent RNA helicase DDX58 (DDX58) | O95786 | Tumor necrosis factor receptor superfamily member EDAR (EDAR) | Q9UNE0 |
| Protein FAM3B (FAM3B) | P58499 | Zinc finger and BTB domain-containing protein 16 (ZBTB16) | Q05516 |

For more details visit www.olink.com/immune-response

### www.olink.com

For research use only. Not for use in diagnostic procedures.
This product includes a license for non-commercial use. Commercial users may require additional licenses. Please contact Olink Proteomics AB for details.
There are no warranties, expressed or implied, which extend beyond this description. Olink Proteomics AB is not liable for property damage, personal injury, or economic loss caused by this product.
Olink® is a registered trademark of Olink Proteomics AB.

© 2017–2022 Olink Proteomics AB. All third party trademarks are the property of their respective owners.

Olink Proteomics, Dag Hammarskjölds väg 52B , SE-752 37 Uppsala, Sweden

1051, v2.0, 2022-06-14

Olink®

Protein assay list

### Olink® Target 96 Inflammation

Product number: 95302

| | |
|---|---|
| Adenosine Deaminase (ADA) | P00813 |
| Artemin (ARTN) | Q5T4W7 |
| Axin-1 (AXIN1) | O15169 |
| Beta-nerve growth factor (Beta-NGF) | P01138 |
| Caspase-8 (CASP-8) | Q14790 |
| C-C motif chemokine 3 (CCL3) | P10147 |
| C-C motif chemokine 4 (CCL4) | P13236 |
| C-C motif chemokine 19 (CCL19) | Q99731 |
| C-C motif chemokine 20 (CCL20) | P78556 |
| C-C motif chemokine 23 (CCL23) | P55773 |
| C-C motif chemokine 25 (CCL25) | O15444 |
| C-C motif chemokine 28 (CCL28) | Q9NRJ3 |
| CD40L receptor (CD40) | P25942 |
| CUB domain-containing protein 1 (CDCP1) | Q9H5V8 |
| C-X-C motif chemokine 1 (CXCL1) | P09341 |
| C-X-C motif chemokine 5 (CXCL5) | P42830 |
| C-X-C motif chemokine 6 (CXCL6) | P80162 |
| C-X-C motif chemokine 9 (CXCL9) | Q07325 |
| C-X-C motif chemokine 10 (CXCL10) | P02778 |
| C-X-C motif chemokine 11 (CXCL11) | O14625 |
| Cystatin D (CST5) | P28325 |
| Delta and Notch-like epidermal growth factor-related receptor (DNER) | Q8NFT8 |
| Eotaxin (CCL11) | P51671 |
| Eukaryotic translation initiation factor 4E-binding protein 1 (4E-BP1) | Q13541 |
| Fibroblast growth factor 21 (FGF-21) | Q9NSA1 |
| Fibroblast growth factor 23 (FGF-23) | Q9GZV9 |
| Fibroblast growth factor 5 (FGF-5) | Q8NF90 |
| Fibroblast growth factor 19 (FGF-19) | O95750 |
| Fms-related tyrosine kinase 3 ligand (Flt3L) | P49771 |
| Fractalkine (CX3CL1) | P78423 |
| Glial cell line-derived neurotrophic factor (GDNF) | P39905 |
| Hepatocyte growth factor (HGF) | P14210 |
| Interferon gamma (IFN-gamma) | P01579 |
| Interleukin-1 alpha (IL-1 alpha) | P01583 |
| Interleukin-2 (IL-2) | P60568 |
| Interleukin-2 receptor subunit beta (IL-2RB) | P14784 |
| Interleukin-4 (IL-4) | P05112 |
| Interleukin-5 (IL5) | P05113 |
| Interleukin-6 (IL6) | P05231 |
| Interleukin-7 (IL-7) | P13232 |
| Interleukin-8 (IL-8) | P10145 |
| Interleukin-10 (IL10) | P22301 |
| Interleukin-10 receptor subunit alpha (IL-10RA) | Q13651 |
| Interleukin-10 receptor subunit beta (IL-10RB) | Q08334 |
| Interleukin-12 subunit beta (IL-12B) | P29460 |
| Interleukin-13 (IL-13) | P35225 |
| Interleukin-15 receptor subunit alpha (IL-15RA) | Q13261 |
| Interleukin-17A (IL-17A) | Q16552 |
| Interleukin-17C (IL-17C) | Q9P0M4 |
| Interleukin-18 (IL-18) | Q14116 |

Table continues on reverse ▶

| | | | |
|---|---|---|---|
| Interleukin-18 receptor 1 (IL-18R1) | Q13478 | Programmed cell death 1 ligand 1 (PD-L1) | Q9NZQ7 |
| Interleukin-20 (IL-20) | Q9NYY1 | Protein S100-A12 (EN-RAGE) | P80511 |
| Interleukin-20 receptor subunit alpha (IL-20RA) | Q9UHF4 | Signaling lymphocytic activation molecule (SLAMF1) | Q13291 |
| Interleukin-22 receptor subunit alpha-1 (IL-22 RA1) | Q8N6P7 | SIR2-like protein 2 (SIRT2) | Q8IXJ6 |
| Interleukin-24 (IL-24) | Q13007 | STAM-binding protein (STAMBP) | O95630 |
| Interleukin-33 (IL-33) | O95760 | Stem cell factor (SCF) | P21583 |
| Latency-associated peptide transforming growth factor beta-1 (LAP TGF-beta-1) | P01137 | Sulfotransferase 1A1 (ST1A1) | P50225 |
| Leukemia inhibitory factor (LIF) | P15018 | T cell surface glycoprotein CD6 isoform (CD6) | Q8WWJ7 |
| Leukemia inhibitory factor receptor (LIF-R) | P42702 | T-cell surface glycoprotein CD5 (CD5) | P06127 |
| Macrophage colony-stimulating factor 1 (CSF-1) | P09603 | T-cell surface glycoprotein CD8 alpha chain (CD8A) | P01732 |
| Matrix metalloproteinase-1 (MMP-1) | P03956 | Thymic stromal lymphopoietin (TSLP) | Q969D9 |
| Matrix metalloproteinase-10 (MMP-10) | P09238 | TNF-beta (TNFB) | P01374 |
| Monocyte chemotactic protein 1 (MCP-1) | P13500 | TNF-related activation-induced cytokine (TRANCE) | O14788 |
| Monocyte chemotactic protein 2 (MCP-2) | P80075 | TNF-related apoptosis-inducing ligand (TRAIL) | P50591 |
| Monocyte chemotactic protein 3 (MCP-3) | P80098 | Transforming growth factor alpha (TGF-alpha) | P01135 |
| Monocyte chemotactic protein 4 (MCP-4) | Q99616 | Tumor necrosis factor (Ligand) superfamily, member 12 (TWEAK) | O43508 |
| Natural killer cell receptor 2B4 (CD244) | Q9BZW8 | Tumor necrosis factor (TNF) | P01375 |
| Neurotrophin-3 (NT-3) | P20783 | Tumor necrosis factor ligand superfamily member 14 (TNFSF14) | O43557 |
| Neurturin (NRTN) | Q99748 | Tumor necrosis factor receptor superfamily member 9 (TNFRSF9) | Q07011 |
| Oncostatin-M (OSM) | P13725 | Urokinase-type plasminogen activator (uPA) | P00749 |
| Osteoprotegerin (OPG) | O00300 | Vascular endothelial growth factor A (VEGF-A) | P15692 |

For more details visit www.olink.com/inflammation

www.olink.com

For research use only. Not for use in diagnostic procedures.
This product includes a license for non-commercial use. Commercial users may require additional licenses. Please contact Olink Proteomics AB for details.
There are no warranties, expressed or implied, which extend beyond this description. Olink Proteomics AB is not liable for property damage, personal injury, or economic loss caused by this product.
Olink® is a registered trademark of Olink Proteomics AB.

© 2017–2022 Olink Proteomics AB. All third party trademarks are the property of their respective owners.

Olink Proteomics, Dag Hammarskjölds väg 52B , SE-752 37 Uppsala, Sweden
1029, v2.0, 2022-06-14

Olink®

Protein assay list

### Olink® Target 96 Metabolism

Product number: 95340

| | |
|---|---|
| Adenosylhomocysteinase (AHCY) | P23526 |
| Adhesion G protein-coupled receptor E2 (ADGRE2) | Q9UHX3 |
| Adhesion G-protein coupled receptor G2 (ADGRG2) | Q8IZP9 |
| Amyloid-like protein 1 (APLP1) | P51693 |
| Angiopoietin-2 (ANGPT2) | O15123 |
| Angiopoietin-related protein 1 (ANGPTL1) | O95841 |
| Angiopoietin-related protein 7 (ANGPTL7) | O43827 |
| Annexin A4 (ANXA4) | P09525 |
| Annexin A11 (ANXA11) | P50995 |
| Appetite-regulating hormone (GHRL) | Q9UBU3 |
| Arginase-1 (ARG1) | P05089 |
| Aromatic-L-amino-acid decarboxylase (DDC) | P20711 |
| B-cell antigen receptor complex-associated protein beta chain (CD79B) | P40259 |
| Cadherin-2 (CDH2) | P19022 |
| Cadherin-related family member 5 (CDHR5) | Q9HBB8 |
| Calsyntenin-2 (CLSTN2) | Q9H4D0 |
| Carbonic anhydrase 13 (CA13) | Q8N1Q1 |
| Catechol O-methyltransferase (COMT) | P21964 |
| Cathepsin O (CTSO) | P43234 |
| CD2-associated protein (CD2AP) | Q9Y5K6 |
| Chordin-like protein 2 (CHRDL2) | Q6WN34 |
| Clusterin-like protein 1 (CLUL1) | Q15846 |
| Coiled-coil domain-containing protein 80 (CCDC80) | Q76M96 |
| Crk-like protein (CRKL) | P46109 |
| C-type lectin domain family 5 member A (CLEC5A) | Q9NY25 |
| CXADR-like membrane protein (CLMP) | Q9H6B4 |
| Diablo homolog, mitochondrial (DIABLO) | Q9NR28 |
| Dihydropteridine reductase (QDPR) | P09417 |
| Dipeptidyl peptidase 2 (DPP7) | Q9UHL4 |
| Disabled homolog 2 (DAB2) | P98082 |
| DNA-(apurinic or apyrimidinic site) lyase (APEX1) | P27695 |
| Ectonucleoside triphosphate diphosphohydrolase 5 (ENTPD5) | O75356 |
| Ectonucleotide pyrophosphatase/phosphodiesterase family member 7 (ENPP7) | Q6UWV6 |
| Eosinophil cationic protein (RNASE3) | P12724 |
| Fc receptor-like protein 1 (FCRL1) | Q96LA6 |
| Fructose-1,6-bisphosphatase 1 (FBP1) | P09467 |
| Galanin peptides (GAL) | P22466 |
| Gamma-enolase (ENO2) | P09104 |
| Glutaredoxin-1 (GLRX) | P35754 |
| GRB2-related adapter protein 2 (GRAP2) | O75791 |
| Hepatoma-derived growth factor (HDGF) | P51858 |
| Inactive tyrosine-protein kinase transmembrane receptor ROR1 (ROR1) | Q01973 |
| Insulin-like growth factor-binding protein-like 1 (IGFBPL1) | Q8WX77 |
| Integrin beta-7 (ITGB7) | P26010 |
| Kallikrein-10 (KLK10) | O43240 |
| Kynurenine-oxoglutarate transaminase 1 (KYAT1) | Q16773 |
| Large proline-rich protein BAG6 (BAG6) | P46379 |
| Leucine-rich repeats and immunoglobulin-like domains protein 1 (LRIG1) | Q96JA1 |
| Leukocyte immunoglobulin-like receptor subfamily A member 5 (LILRA5) | A6NI73 |

Table continues on reverse ▶

| | |
|---|---|
| Low-density lipoprotein receptor-related protein 11 (LRP11) | Q86VZ4 |
| Lysophosphatidic acid phosphatase type 6 (ACP6) | Q9NPH0 |
| Meprin A subunit beta (MEP1B) | Q16820 |
| Meteorin-like protein (METRNL) | Q641Q3 |
| Multiple coagulation factor deficiency protein 2 (MCFD2) | Q8NI22 |
| NAD kinase (NADK) | O95544 |
| Nectin-2 (NECTIN2) | Q92692 |
| Neural proliferation differentiation and control protein 1 (NPDC1) | Q9NQX5 |
| Neuronal pentraxin receptor (NPTXR) | O95502 |
| Nodal modulator 1 (NOMO1) | Q15155 |
| N-terminal prohormone brain natriuretic peptide (NT-proBNP) | NA |
| Paired immunoglobulin-like type 2 receptor beta (PILRB) | Q9UKJ0 |
| Peptidyl-prolyl cis-trans isomerase FKBP4 (FKBP4) | Q02790 |
| Phosphoprotein associated with glycosphingolipid-enriched microdomains 1 (PAG1) | Q9NWQ8 |
| Pro-cathepsin H (CTSH) | P09668 |
| Protein FAM3C (FAM3C) | Q92520 |
| Protein phosphatase inhibitor 2 (PPP1R2) | P41236 |
| Protein S100-P (S100P) | P25815 |
| Regenerating islet-derived protein 4 (REG4) | Q9BYZ8 |
| Reticulon-4 receptor (RTN4R) | Q9BZR6 |
| Retinal dehydrogenase 1 (ALDH1A1) | P00352 |
| Ribosyldihydronicotinamide dehydrogenase [quinone] (NQO2) | P16083 |
| Scavenger receptor cysteine-rich domain-containing group B protein (SSC4D) | Q8WTU2 |
| Sclerostin (SOST) | Q9BQB4 |
| Semaphorin-3F (SEMA3F) | Q13275 |
| Serpin B6 (SERPINB6) | P35237 |
| Serpin B8 (SERPINB8) | P50452 |
| Sialic acid-binding Ig-like lectin 7 (SIGLEC7) | Q9Y286 |
| Sialomucin core protein 24 (CD164) | Q04900 |
| Soluble calcium-activated nucleotidase 1 (CANT1) | Q8WVQ1 |
| Sulfatase-modifying factor 2 (SUMF2) | Q8NBJ7 |
| Synaptosomal-associated protein 23 (SNAP23) | O00161 |
| Syndecan-4 (SDC4) | P31431 |
| T-cell surface glycoprotein CD1c (CD1C) | P29017 |
| Thimet oligopeptidase (THOP1) | P52888 |
| Thioredoxin domain-containing protein 5 (TXNDC5) | Q8NBS9 |
| Thymidine phosphorylase (TYMP) | P19971 |
| Thyrotropin subunit beta (TSHB) | P01222 |
| Trefoil factor 2 (TFF2) | Q03403 |
| Tubulointerstitial nephritis antigen-like (TINAGL1) | Q9GZM7 |
| Tyrosine-protein kinase receptor TYRO3 (TYRO3) | Q06418 |
| Ubiquitin carboxyl-terminal hydrolase 8 (USP8) | P40818 |
| Versican core protein (VCAN) | P13611 |

For more details visit www.olink.com/metabolism

www.olink.com

For research use only. Not for use in diagnostic procedures.
This product includes a license for non-commercial use. Commercial users may require additional licenses. Please contact Olink Proteomics AB for details.
There are no warranties, expressed or implied, which extend beyond this description. Olink Proteomics AB is not liable for property damage, personal injury, or economic loss caused by this product.
Olink® is a registered trademark of Olink Proteomics AB.

© 2017–2022 Olink Proteomics AB. All third party trademarks are the property of their respective owners.
Olink Proteomics, Dag Hammarskjölds väg 52B , SE-752 37 Uppsala, Sweden
1053, v2.0, 2022-06-14

Olink®

Protein assay list

### Olink® Target 96 Neuro Exploratory

Product number: 95391

| | |
|---|---|
| 6-pyruvoyl tetrahydrobiopterin synthase (PTS) | Q03393 |
| Adhesion G protein-coupled receptor B3 (ADGRB3) | O60242 |
| Alanyl-tRNA editing protein Aarsd1 (AARSD1) | Q9BTE6 |
| Alpha-(1,6)-fucosyltransferase (FUT8) | Q9BYC5 |
| Amiloride-sensitive amine oxidase [copper-containing] (AOC1) | P19801 |
| Annexin A10 (ANXA10) | Q9UJ72 |
| Asialoglycoprotein receptor 1 (ASGR1) | P07306 |
| Beta-defensin 4A (DEFB4A) | O15263 |
| Beta-klotho (KLB) | Q86Z14 |
| Bis(5'-adenosyl)-triphosphatase (FHIT) | P49789 |
| Bone marrow stromal antigen 2 (BST2) | Q10589 |
| Cadherin-15 (CDH15) | P55291 |
| Cadherin-17 (CDH17) | Q12864 |
| Calcineurin subunit B type 1 (PPP3R1) | P63098 |
| Calcium-regulated heat-stable protein 1 (CARHSP1) | Q9Y2V2 |
| Calsyntenin-1 (CLSTN1) | O94985 |
| Carcinoembryonic antigen-related cell adhesion molecule 3 (CEACAM3) | P40198 |
| Cardiotrophin-1 (CTF1) | Q16619 |
| C-C motif chemokine 27 (CCL27) | Q9Y4X3 |
| CD302 antigen (CD302) | Q8IX05 |
| CD63 antigen (CD63) | P08962 |
| Centrin-2 (CETN2) | P41208 |
| Ceramide transfer protein (CERT1) | Q9Y5P4 |
| Cysteine-rich protein 2 (CRIP2) | P52943 |
| Death domain-containing protein CRADD (CRADD) | P78560 |
| Desmoglein-3 (DSG3) | P32926 |
| Dipeptidase 1 (DPEP1) | P16444 |
| Dipeptidase 2 (DPEP2) | Q9H4A9 |
| Disintegrin and metalloproteinase domain-containing protein 15 (ADAM15) | Q13444 |
| Dual specificity protein phosphatase 3 (DUSP3) | P51452 |
| E3 ubiquitin-protein ligase RNF31 (RNF31) | Q96EP0 |
| Endoplasmin (HSP90B1) | P14625 |
| Endothelin-converting enzyme 1 (ECE1) | P42892 |
| Ephrin type-A receptor 10 (EPHA10) | Q5JZY3 |
| Eukaryotic translation initiation factor 4B (EIF4B) | P23588 |
| Fibroblast growth factor receptor 2 (FGFR2) | P21802 |
| Gamma-interferon-inducible lysosomal thiol reductase (IFI30) | P13284 |
| Gamma-synuclein (SNCG) | O76070 |
| Glutathione hydrolase 5 proenzyme (GGT5) | P36269 |
| Glutathione S-transferase P (GSTP1) | P09211 |
| Glycodelin (PAEP) | P09466 |
| Group 10 secretory phospholipase A2 (PLA2G10) | O15496 |
| Guanylate-binding protein 2 (GBP2) | P32456 |
| Heme oxygenase 2 (HMOX2) | P30519 |
| Immunoglobulin alpha Fc receptor (FCAR) | P24071 |
| Immunoglobulin superfamily containing leucine-rich repeat protein 2 (ISLR2) | Q6UXK2 |
| Inhibitor of growth protein 1 (ING1) | Q9UK53 |
| Inositol monophosphatase 1 (IMPA1) | P29218 |
| Integrin-linked kinase-associated serine/threonine phosphatase 2C (ILKAP) | Q9H0C8 |
| Interferon lambda-1 (IFNL1) | Q8IU54 |
| Interleukin-15 (IL15) | P40933 |

Table continues on reverse ▶

| | | | |
|---|---|---|---|
| Interleukin-3 receptor subunit alpha (IL3RA) | P26951 | Pregnancy-specific beta-1-glycoprotein 1 (PSG1) | P11464 |
| Interleukin-32 (IL32) | P24001 | Proepiregulin (EREG) | O14944 |
| KIF1-binding protein (KIF1BP) | Q96EK5 | Proline-rich AKT1 substrate 1 (AKT1S1) | Q96B36 |
| Killer cell immunoglobulin-like receptor 2DL3 (KIR2DL3) | P43628 | Proteasome activator complex subunit 1 (PSME1) | Q06323 |
| Kin of IRRE-like protein 2 (KIRREL2) | Q6UWL6 | Protein ABHD14B (ABHD14B) | Q96IU4 |
| Latent-transforming growth factor beta-binding protein 3 (LTBP3) | Q9NS15 | Protein NDRG1 (NDRG1) | Q92597 |
| Leptin receptor (LEPR) | P48357 | Ribokinase (RBKS) | Q9H477 |
| Mitotic spindle assembly checkpoint protein MAD1 (MAD1L1) | Q9Y6D9 | Ribosomal protein S6 kinase beta-1 (RPS6KB1) | P23443 |
| Myeloid cell surface antigen CD33 (CD33) | P20138 | Secreted frizzled-related protein 1 (SFRP1) | Q8N474 |
| N-alpha-acetyltransferase 10 (NAA10) | P41227 | Signal recognition particle 14 kDa protein (SRP14) | P37108 |
| NEDD4-like E3 ubiquitin-protein ligase WWP2 (WWP2) | O00308 | SPARC-related modular calcium-binding protein 1 (SMOC1) | Q9H4F8 |
| NEDD8-conjugating enzyme UBE2F (UBE2F) | Q969M7 | Teratocarcinoma-derived growth factor 1 (TDGF1) | P13385 |
| Neurexophilin-1 (NXPH1) | P58417 | Transmembrane glycoprotein NMB (GPNMB) | Q14956 |
| Neurofilament light polypeptide (NEFL) | P07196 | Tubulin polymerization-promoting protein family member 3 (TPPP3) | Q9BW30 |
| Nucleophosmin (NPM1) | P06748 | Tubulin-folding cofactor B (TBCB) | Q99426 |
| Peptidyl-prolyl cis-trans isomerase FKBP5 (FKBP5) | Q13451 | Tumor necrosis factor receptor superfamily member 13C (TNFRSF13C) | Q96RJ3 |
| Peptidyl-prolyl cis-trans isomerase FKBP7 (FKBP7) | Q9Y680 | Tyrosine-protein phosphatase non-receptor type 1 (PTPN1) | P18031 |
| Phosphoethanolamine/phosphocholine phosphatase (PHOSPHO1) | Q8TCT1 | V-set and transmembrane domain-containing protein 1 (VSTM1) | Q6UX27 |
| Phosphomevalonate kinase (PMVK) | Q15126 | V-type proton ATPase subunit F (ATP6V1F) | Q16864 |
| Phosphoribosyltransferase domain-containing protein 1 (PRTFDC1) | Q9NRG1 | Zinc finger protein Helios (IKZF2) | Q9UKS7 |
| Prefoldin subunit 2 (PFDN2) | Q9UHV9 | | |

For more details visit www.olink.com/neuro-exploratory

### www.olink.com

For research use only. Not for use in diagnostic procedures.
This product includes a license for non-commercial use. Commercial users may require additional licenses. Please contact Olink Proteomics AB for details.
There are no warranties, expressed or implied, which extend beyond this description. Olink Proteomics AB is not liable for property damage, personal injury, or economic loss caused by this product.
Olink® is a registered trademark of Olink Proteomics AB.

© 2017–2022 Olink Proteomics AB. All third party trademarks are the property of their respective owners.

Olink Proteomics, Dag Hammarskjölds väg 52B , SE-752 37 Uppsala, Sweden
1100, v2.0, 2022-08-16

Olink®

Protein assay list

### Olink® Target 96 Neurology

Product number: 95801

| | | | |
|---|---|---|---|
| ADP-ribosyl cyclase/cyclic ADP-ribose hydrolase 1 (CD38) | P28907 | Ephrin type-B receptor 6 (EPHB6) | O15197 |
| Alpha-2-macroglobulin receptor-associated protein (Alpha-2-MRAP) | P30533 | Ephrin-A4 (EFNA4) | P52798 |
| | | Epithelial discoidin domain-containing receptor 1 (DDR1) | Q08345 |
| BDNF/NT-3 growth factors receptor (NTRK2) | Q16620 | Ezrin (EZR) | P15311 |
| Beta-nerve growth factor (Beta-NGF) | P01138 | Fc receptor-like protein 2 (FcRL2) | Q96LA5 |
| Bone morphogenetic protein 4 (BMP-4) | P12644 | | |
| Brevican core protein (BCAN) | Q96GW7 | Galectin-8 (gal-8) | O00214 |
| Brorin (VWC2) | Q2TAL6 | GDNF family receptor alpha-1 (GFR-alpha-1) | P56159 |
| Cadherin-3 (CDH3) | P22223 | GDNF family receptor alpha-3 (GDNFR-alpha-3) | O60609 |
| Cadherin-6 (CDH6) | P55285 | Glial cell line-derived neurotrophic factor (GDNF) | P39905 |
| | | Glypican-5 (GCP5) | P78333 |
| Carboxypeptidase A2 (CPA2) | P48052 | Granulocyte Colony-Stimulating Factor (G-CSF) | P09919 |
| Carboxypeptidase M (CPM) | P14384 | Granulocyte-macrophage colony-stimulating factor receptor subunit alpha (GM-CSF-R-alpha) | P15509 |
| Cathepsin S (CTSS) | P25774 | | |
| Cell adhesion molecule 3 (CADM3) | Q8N126 | Granzyme A (GZMA) | P12544 |
| Cell surface glycoprotein CD200 receptor 1 (CD200R1) | Q8TD46 | Growth/differentiation factor 8 (GDF-8) | O14793 |
| | | Hydroxyacylglutathione hydrolase, mitochondrial (HAGH) | Q16775 |
| CMRF35-like molecule 1 (CLM-1) | Q8TDQ1 | Interleukin-5 receptor subunit alpha (IL-5R-alpha) | Q01344 |
| CMRF35-like molecule 6 (CLM-6) | Q08708 | Interleukin-12 (IL-12) | P29460, P29459 |
| Contactin-5 (CNTN5) | O94779 | | |
| C-type lectin domain family 1 member B (CLEC1B) | Q9P126 | Junctional adhesion molecule B (JAM-B) | P57087 |
| C-type lectin domain family 10 member A (CLEC10A) | Q8IUN9 | Kynureninase (KYNU) | Q16719 |
| Cytotoxic and regulatory T-cell molecule (CRTAM) | O95727 | Latexin (LXN) | Q9BS40 |
| Dickkopf-related protein 4 (Dkk-4) | Q9UBT3 | Layilin (LAYN) | Q6UX15 |
| Dipeptidyl peptidase 1 (CTSC) | P53634 | Leucine-rich repeat transmembrane protein FLRT2 (FLRT2) | O43155 |
| Disintegrin and metalloproteinase domain-containing protein 22 (ADAM 22) | Q9P0K1 | Leukocyte-associated immunoglobulin-like receptor 2 (LAIR-2) | Q6ISS4 |
| Disintegrin and metalloproteinase domain-containing protein 23 (ADAM 23) | O75077 | Linker for activation of T-cells family member 1 (LAT) | O43561 |
| Draxin (DRAXIN) | Q8NBI3 | Lysosome membrane protein 2 (SCARB2) | Q14108 |

Table continues on reverse ▶

| | | | |
|---|---|---|---|
| Macrophage scavenger receptor types I and II (MSR1) | P21757 | Protogenin (PRTG) | Q2VWP7 |
| MAM domain-containing glycosylphosphatidylinositol anchor protein 1 (MDGA1) | Q8NFP4 | Repulsive guidance molecule A (RGMA) | Q96B86 |
| | | RGM domain family member B (RGMB) | Q6NW40 |
| Matrilin-3 (MATN3) | O15232 | | |
| Mesencephalic astrocyte-derived neurotrophic factor (MANF) | P55145 | Roundabout homolog 2 (ROBO2) | Q9HCK4 |
| Microtubule-associated protein tau (MAPT) | P10636 | R-spondin-1 (RSPO1) | Q2MKA7 |
| N-acylethanolamine-hydrolyzing acid amidase (NAAA) | Q02083 | Scavenger receptor class A member 5 (SCARA5) | Q6ZMJ2 |
| Neprilysin (NEP) | P08473 | Scavenger receptor class F member 2 (SCARF2) | Q96GP6 |
| Netrin receptor UNC5C (UNC5C) | O95185 | Secreted frizzled-related protein 3 (sFRP-3) | Q92765 |
| Neuroblastoma suppressor of tumorigenicity 1 (NBL1) | P41271 | Serine/threonine-protein kinase receptor R3 (SKR3) | P37023 |
| Neurocan core protein (NCAN) | O14594 | Sialic acid-binding Ig-like lectin 9 (Siglec-9) | Q9Y336 |
| Neuronal cell adhesion molecule (Nr-CAM) | Q92823 | Sialoadhesin (SIGLEC1) | Q9BZZ2 |
| Neuropilin-2 (NRP2) | O60462 | SPARC-related modular calcium-binding protein 2 (SMOC2) | Q9H3U7 |
| Neutral ceramidase (N-CDase) | Q9NR71 | Sphingomyelin phosphodiesterase (SMPD1) | P17405 |
| Nicotinamide/nicotinic acid mononucleotide adenylyltransferase 1 (NMNAT1) | Q9HAN9 | Tenascin-R (TN-R) | Q92752 |
| | | Testican-1 (SPOCK1) | Q08629 |
| NKG2D ligand 2 (N2DL-2) | Q9BZM5 | | |
| NT-3 growth factor receptor (NTRK3) | Q16288 | Thy-1 membrane glycoprotein (THY 1) | P04216 |
| OX-2 membrane glycoprotein (CD200) | P41217 | Transmembrane protease serine 5 (TMPRSS5) | Q9H3S3 |
| Platelet-derived growth factor receptor alpha (PDGF-R-alpha) | P16234 | Tumor necrosis factor receptor superfamily member 12A (TNFRSF12A) | Q9NP84 |
| Plexin-B1 (PLXNB1) | O43157 | Tumor necrosis factor receptor superfamily member 21 (TNFRSF21) | O75509 |
| Plexin-B3 (PLXNB3) | Q9ULL4 | Tumor necrosis factor receptor superfamily member 27 (EDA2R) | Q9HAV5 |
| Poliovirus receptor (PVR) | P15151 | WAP, Kazal, immunoglobulin, Kunitz and NTR domain-containing protein 1 (WFIKKN1) | Q96NZ8 |

For more details visit www.olink.com/neurology

www.olink.com

For research use only. Not for use in diagnostic procedures.
This product includes a license for non-commercial use. Commercial users may require additional licenses. Please contact Olink Proteomics AB for details.
There are no warranties, expressed or implied, which extend beyond this description. Olink Proteomics AB is not liable for property damage, personal injury, or economic loss caused by this product.
Olink® is a registered trademark of Olink Proteomics AB.

© 2017–2022 Olink Proteomics AB. All third party trademarks are the property of their respective owners.
Olink Proteomics, Dag Hammarskjölds väg 52B , SE-752 37 Uppsala, Sweden
1035, v2.0, 2022-06-16

Olink®

Protein assay list

### Olink® Target 96 Oncology II

Product number: 95700

| | | | |
|---|---|---|---|
| 5'-nucleotidase (5'-NT) | P21589 | Fc receptor-like B (FCRLB) | Q6BAA4 |
| A disintegrin and metalloproteinase with thrombospondin motifs 15 (ADAM-TS 15) | Q8TE58 | Fibroblast growth factor-binding protein 1 (FGF-BP1) | Q14512 |
| Alpha-taxilin (TXLNA) | P40222 | Folate receptor alpha (FR-alpha) | P15328 |
| Amphiregulin (AREG) | P15514 | Folate receptor gamma (FR-gamma) | P41439 |
| Annexin A1 (ANXA1) | P04083 | Furin (FUR) | P09958 |
| Carbonic anhydrase IX (CAIX) | Q16790 | Galectin-1 (Gal-1) | P09382 |
| Carboxypeptidase E (CPE) | P16870 | Glypican-1 (GPC1) | P35052 |
| Carcinoembryonic antigen (CEA) | P06731 | Granzyme B (GZMB) | P10144 |
| Carcinoembryonic antigen-related cell adhesion molecule 1 (CEACAM1) | P13688 | Granzyme H (GZMH) | P20718 |
| Cathepsin L2 (CTSV) | O60911 | Hepatocyte growth factor (HGF) | P14210 |
| CD27 antigen (CD27) | P26842 | ICOS ligand (ICOSLG) | O75144 |
| CD160 antigen (CD160) | O95971 | Insulin-like growth factor 1 receptor (IGF1R) | P08069 |
| CD48 antigen (CD48) | P09326 | Integrin alpha-V (ITGAV) | P06756 |
| CD70 antigen (CD70) | P32970 | Integrin beta-5 (ITGB5) | P18084 |
| Cornulin (CRNN) | Q9UBG3 | Interferon gamma receptor 1 (IFN-gamma-R1) | P15260 |
| C-type lectin domain family 4 member K (CD207) | Q9UJ71 | Interleukin-6 (IL6) | P05231 |
| C-X-C motif chemokine 13 (CXCL13) | O43927 | Kallikrein-8 (hK8) | O60259 |
| Cyclin-dependent kinase inhibitor 1 (CDKN1A) | P38936 | Kallikrein-11 (hK11) | Q9UBX7 |
| Delta-like protein 1 (DLL1) | O00548 | Kallikrein-13 (KLK13) | Q9UKR3 |
| Disintegrin and metalloproteinase domain-containing protein 8 (ADAM8) | P78325 | Kallikrein-14 (hK14) | Q9P0G3 |
| | | Ly6/PLAUR domain-containing protein 3 (LYPD3) | O95274 |
| Endothelial cell-specific molecule 1 (ESM-1) | Q9NQ30 | Melanoma-derived growth regulatory protein (MIA) | Q16674 |
| Ephrin type-A receptor 2 (EPHA2) | P29317 | Mesothelin (MSLN) | Q13421 |
| Fas antigen ligand (FasL) | P48023 | Methionine aminopeptidase 2 (MetAP 2) | P50579 |
| FAS-associated death domain protein (FADD) | Q13158 | MHC class I polypeptide-related sequence A/B (MIC-A/B) | Q29983, Q29980 |

Table continues on reverse ▶

| | |
|---|---|
| Midkine (MK) | P21741 |
| Mothers against decapentaplegic homolog 5 (MAD homolog 5) | Q99717 |
| Mucin-16 (MUC-16) | Q8WXI7 |
| Nectin-4 (PVRL4) | Q96NY8 |
| Pancreatic prohormone (PPY) | P01298 |
| Podocalyxin (PODXL) | O00592 |
| Pro-epidermal growth factor (EGF) | P01133 |
| Protein CYR61 (CYR61) | O00622 |
| Protein S100-A11 (S100A11) | P31949 |
| Protein S100-A4 (S100A4) | P26447 |
| Proto-oncogene tyrosine-protein kinase receptor Ret (RET) | P07949 |
| Receptor tyrosine-protein kinase erbB-2 (ErbB2/HER2) | P04626 |
| Receptor tyrosine-protein kinase erbB-3 (ErbB3/HER3) | P21860 |
| Receptor tyrosine-protein kinase erbB-4 (ErbB4/HER4) | Q15303 |
| R-spondin-3 (RSPO3) | Q9BXY4 |
| Secretory carrier-associated membrane protein 3 (SCAMP3) | O14828 |
| Seizure 6-like protein (SEZ6L) | Q9BYH1 |
| SPARC (SPARC) | P09486 |
| Stem cell factor (SCF) | P21583 |
| Syndecan-1 (SYND1) | P18827 |
| T-cell leukemia / lymphoma protein 1A (TCL1A) | P56279 |
| TGF-beta receptor type-2 (TGFR-2) | P37173 |
| Tissue factor pathway inhibitor 2 (TFPI-2) | P48307 |
| T-lymphocyte surface antigen Ly-9 (LY9) | Q9HBG7 |
| TNF-related apoptosis-inducing ligand (TRAIL) | P50591 |
| Toll-like receptor 3 (TLR3) | O15455 |
| Transforming growth factor alpha (TGF-alpha) | P01135 |
| Transmembrane glycoprotein NMB (GPNMB) | Q14956 |
| Tumor necrosis factor ligand superfamily member 13 (TNFSF13) | O75888 |
| Tumor necrosis factor receptor superfamily member 4 (TNFRSF4) | P43489 |
| Tumor necrosis factor receptor superfamily member 6B (TNFRSF6B) | O95407 |
| Tumor necrosis factor receptor superfamily member 19 (TNFRSF19) | Q9NS68 |
| Tyrosine-protein kinase ABL1 (ABL1) | P00519 |
| Tyrosine-protein kinase Lyn (LYN) | P07948 |
| WAP four-disulfide core domain protein 2 (WFDC2) | Q14508 |
| Vascular endothelial growth factor A (VEGF-A) | P15692 |
| Vascular endothelial growth factor receptor 2 (VEGFR-2) | P35968 |
| Vascular endothelial growth factor receptor 3 (VEGFR-3) | P35916 |
| VEGF-co regulated chemokine 1 (CXL17) | Q6UXB2 |
| Vimentin (VIM) | P08670 |
| Wnt inhibitory factor 1 (WIF-1) | Q9Y5W5 |
| WNT1-inducible-signaling pathway protein 1 (WISP-1) | O95388 |
| Xaa-Pro aminopeptidase 2 (XPNPEP2) | O43895 |

For more details visit www.olink.com/onc2

### www.olink.com

For research use only. Not for use in diagnostic procedures.
This product includes a license for non-commercial use. Commercial users may require additional licenses. Please contact Olink Proteomics AB for details.
There are no warranties, expressed or implied, which extend beyond this description. Olink Proteomics AB is not liable for property damage, personal injury, or economic loss caused by this product.
Olink® is a registered trademark of Olink Proteomics AB.

© 2017–2022 Olink Proteomics AB. All third party trademarks are the property of their respective owners.

Olink Proteomics, Dag Hammarskjölds väg 52B , SE-752 37 Uppsala, Sweden

1032, v2.0, 2022-06-14

Protein assay list

### Olink® Target 96 Organ Damage

Product number: 95331

| | |
|---|---|
| 5'-AMP-activated protein kinase subunit beta-1 (PRKAB1) | Q9Y478 |
| Adhesion G-protein coupled receptor G1 (ADGRG1) | Q9Y653 |
| Aldehyde dehydrogenase, dimeric NADP-preferring (ALDH3A1) | P30838 |
| Anterior gradient protein 2 homolog (AGR2) | O95994 |
| Apoptosis-inducing factor 1, mitochondrial (AIFM1) | O95831 |
| B-cell scaffold protein with ankyrin repeats (BANK1) | Q8NDB2 |
| BH3-interacting domain death agonist (BID) | P55957 |
| BMP and activin membrane-bound inhibitor homolog (BAMBI) | Q13145 |
| Calcitonin (CALCA) | P01258 |
| Calreticulin (CALR) | P27797 |
| Carbonic anhydrase 12 (CA12) | O43570 |
| Carbonic anhydrase 14 (CA14) | Q9ULX7 |
| Casein kinase I isoform delta (CSNK1D) | P48730 |
| Claspin (CLSPN) | Q9HAW4 |
| CMP-N-acetylneuraminate-beta-galactosamide-alpha-2,3-sialyltransferase 1 (ST3GAL1) | Q11201 |
| Cocaine esterase (CES2) | O00748 |
| Contactin-2 (CNTN2) | Q02246 |
| Corticoliberin (CRH) | P06850 |
| C-type lectin domain family 1 member A (CLEC1A) | Q8NC01 |
| C-type natriuretic peptide (NPPC) | P23582 |
| Desmoglein-4 (DSG4) | Q86SJ6 |
| Dipeptidyl aminopeptidase-like protein 6 (DPP6) | P42658 |
| DNA topoisomerase 2-beta (TOP2B) | Q02880 |
| Ectonucleoside triphosphate diphosphohydrolase 2 (ENTPD2) | Q9Y5L3 |
| Ectonucleoside triphosphate diphosphohydrolase 6 (ENTPD6) | O75354 |
| EGF-like repeat and discoidin I-like domain-containing protein 3 (EDIL3) | O43854 |
| Enteropeptidase (TMPRSS15) | P98073 |
| Epidermal growth factor-like protein 7 (EGFL7) | Q9UHF1 |
| Erbin (ERBIN) | Q96RT1 |
| Erythropoietin (EPO) | P01588 |
| Fatty acid-binding protein 9 (FABP9) | Q0Z7S8 |
| Forkhead box protein O1 (FOXO1) | Q12778 |
| Fructose-2,6-bisphosphatase TIGAR (TIGAR) | Q9NQ88 |
| Hematopoietic prostaglandin D synthase (HPGDS) | O60760 |
| Inactive tyrosine-protein kinase 7 (PTK7) | Q13308 |
| Integrin beta-1-binding protein 1 (ITGB1BP1) | O14713 |
| Interferon-inducible double-stranded RNA-dependent protein kinase activator A (PRKRA) | O75569 |
| Kidney Injury Molecule (KIM1) | Q96D42 |
| Killer cell immunoglobulin-like receptor 3DL1 (KIR3DL1) | P43629 |
| Leukotriene A-4 hydrolase (LTA4H) | P09960 |
| Linker for activation of T-cells family member 2 (LAT2) | Q9GZY6 |
| Lutropin subunit beta (LHB) | P01229 |
| Macrophage erythroblast attacher (MAEA) | Q7L5Y9 |
| Macrophage-capping protein (CAPG) | P40121 |
| Melanoma-associated antigen D1 (MAGED1) | Q9Y5V3 |
| Methionine aminopeptidase 1 (METAP1) | P53582 |
| Mevalonate kinase (MVK) | Q03426 |
| Mitogen-activated protein kinase kinase kinase kinase 5 (MAP4K5) | Q9Y4K4 |
| Mothers against decapentaplegic homolog 1 (SMAD1) | Q15797 |
| NAD-dependent protein deacylase sirtuin-5, mitochondrial (SIRT5) | Q9NXA8 |
| NEDD8 ultimate buster 1 (NUB1) | Q9Y5A7 |
| Neutrophil cytosol factor 2 (NCF2) | P19878 |

Table continues on reverse ▶

| | | | |
|---|---|---|---|
| Nibrin (NBN) | O60934 | Protein max (MAX) | P61244 |
| Nitric oxide synthase, endothelial (NOS3) | P29474 | Protein phosphatase 1B (PPM1B) | O75688 |
| Nucleobindin-2 (NUCB2) | P80303 | [Pyruvate dehydrogenase [acetyl-transferring]]-phosphatase 1, mitochondrial (PDP1) | Q9P0J1 |
| Parvalbumin alpha (PVALB) | P20472 | Ras association domain-containing protein 2 (RASSF2) | P50749 |
| Paxillin (PXN) | P49023 | Ras GTPase-activating protein 1 (RASA1) | P20936 |
| Peptidyl-prolyl cis-trans isomerase FKBP1B (FKBP1B) | P68106 | Receptor-type tyrosine-protein phosphatase eta (PTPRJ) | Q12913 |
| Perilipin-1 (PLIN1) | O60240 | Renin receptor (ATP6AP2) | O75787 |
| Phosphatidylinositol 3,4,5-trisphosphate 5-phosphatase 2 (INPPL1) | O15357 | REST corepressor 1 (RCOR1) | Q9UKL0 |
| Placenta growth factor (PGF) | P49763 | Retinoic acid receptor responder protein 1 (RARRES1) | P49788 |
| Platelet-derived growth factor C (PDGFC) | Q9NRA1 | Ribonucleoside-diphosphate reductase subunit M2 B (RRM2B) | Q7LG56 |
| Pleiotrophin (PTN) | P21246 | Serpin A9 (SERPINA9) | Q86WD7 |
| Plexin domain-containing protein 1 (PLXDC1) | Q8IUK5 | Serum paraoxonase/arylesterase 2 (PON2) | Q15165 |
| Polypeptide N-acetylgalactosaminyltransferase 10 (GALNT10) | Q86SR1 | Syntaxin-8 (STX8) | Q9UNK0 |
| Probetacellulin (BTC) | P35070 | Syntaxin-binding protein 3 (STXBP3) | O00186 |
| Programmed cell death protein 1 (PDCD1) | Q15116 | Troponin I, cardiac muscle (TNNI3) | P19429 |
| Prolow-density lipoprotein receptor-related protein 1 (LRP1) | Q07954 | Tyrosine-protein kinase Fes/Fps (FES) | P07332 |
| Proteasome subunit alpha type-1 (PSMA1) | P25786 | Tyrosine-protein kinase Fgr (FGR) | P09769 |
| Protein amnionless (AMN) | Q9BXJ7 | Tyrosine-protein kinase Yes (YES1) | P07947 |
| Protein enabled homolog (ENAH) | Q8N8S7 | Vasohibin-1 (VASH1) | Q7L8A9 |
| Protein fosB (FOSB) | P53539 | Wiskott-Aldrich syndrome protein (WAS) | P42768 |

For more details visit www.olink.com/organ-damage

www.olink.com

For research use only. Not for use in diagnostic procedures.
This product includes a license for non-commercial use. Commercial users may require additional licenses. Please contact Olink Proteomics AB for details.
There are no warranties, expressed or implied, which extend beyond this description. Olink Proteomics AB is not liable for property damage, personal injury, or economic loss caused by this product.
Olink® is a registered trademark of Olink Proteomics AB.

© 2017–2022 Olink Proteomics AB. All third party trademarks are the property of their respective owners.

Olink Proteomics, Dag Hammarskjölds väg 52B , SE-752 37 Uppsala, Sweden

1052, v2.0, 2022-06-14
